# A mathematical model for pertussis transmission and vaccination

**DOI:** 10.64898/2026.03.16.26348473

**Authors:** Rachel A. Hounsell, Jared Norman, Rudzani Muloiwa, Sheetal P. Silal

## Abstract

Pertussis remains an endemic and periodically resurgent vaccine-preventable disease despite long-standing immunisation programmes, reflecting complex interactions between transmission, waning immunity, vaccination history, and heterogeneous clinical presentation. We present a comprehensive age-structured mathematical model of pertussis transmission that explicitly represents infection heterogeneity, immunity dynamics, and detailed vaccination schedules across the life course. The model stratifies the population into 56 age groups and 29 epidemiological states, capturing four distinct infection types that differ by severity, symptoms, and infectiousness, including asymptomatic infection. Both naturally acquired and vaccine-derived immunity are modelled as non-lifelong, incorporating waning, partial protection, reinfection, and immune boosting following exposure without transmissible infection. Vaccination is represented at high resolution, including dose-specific primary series vaccination, booster doses in early childhood, childhood, and adolescence, and maternal immunisation during pregnancy, with differentiation between whole-cell and acellular pertussis vaccine formulations and historical changes in vaccine use and coverage. Periodicity and stochasticity are incorporated to reproduce observed multi-year epidemic cycles. A global sensitivity analysis using Latin hypercube sampling and partial rank correlation coefficients identifies immunity waning rates, immune boosting, and recovery from severe infection as key drivers of modelled incidence, mortality, and population protection. By integrating detailed immune processes with realistic vaccination histories, this model provides a flexible framework for evaluating pertussis epidemiology and assessing the population-level impact of alternative vaccination strategies, including booster and maternal immunisation policies.

## 1. Introduction

### 1.1 Biology and pathology of pertussis

Pertussis, commonly known as whooping cough, is a highly contagious, vaccine-preventable respiratory disease caused primarily by the bacterium *Bordetella pertussis*, with *B. parapertussis* causing a milder form of the disease [1,2]. It is transmitted via respiratory droplets. Despite the availability of effective vaccines, pertussis remains a serious public health challenge, causing substantial morbidity and mortality worldwide, particularly among infants under one year of age who are too young to be fully vaccinated [1,2].

*B. pertussis* expresses numerous virulence factors that facilitate colonisation, spread, and resistance to host immunity, contributing to its pathogenicity and the unique disease profile it causes [1,3]. The disease is characterised by paroxysmal coughing fits followed by a distinctive inspiratory “whoop” and posttussive vomiting, although atypical presentations can occur, especially in adults and previously vaccinated individuals. Pertussis is particularly severe in infants under three months, pregnant women, and the elderly, with complications such as pneumonia, seizures, and encephalopathy being most serious in young infants [4]. In low-and middle-income countries (LMIC), the average case fatality rate for pertussis in infants aged <1 year is almost 4% and for children aged 1−4 years is 1% [2]. Older individuals have less severe disease and fewer complications, but substantial economic costs are associated with unrecognised infection in these individuals, who also serve as an important source of infection for non-immune infants [5].

Detection of pertussis can be challenging due to its variable and atypical presentations, especially in previously vaccinated individuals, necessitating the use of diagnostic methods such as polymerase chain reaction (PCR) for accurate identification. Treatment primarily involves macrolide antibiotics such as erythromycin and azithromycin, which can reduce transmission if administered early [4]. Despite advances in diagnostics and treatment, broad vaccination coverage is essential to control the spread of *B. pertussis* and reduce the disease’s circulation [3].

### 1.2 Epidemiology of pertussis

Prior to the availability of pertussis vaccines in the 1950s, pertussis was one of the most common diseases in children <5 years of age, with the highest case-fatality ratios in infancy. After the introduction of the pertussis vaccine into the Expanded Programme on Immunisation (EPI), the number of pertussis cases and deaths in children declined significantly [6]. The Global Burden of Diseases (GBD) study estimated the global burden of pertussis in 2019 was 19.5 million cases, down 41% from 33 million estimated cases in 1990 [7,8]. However, the true burden of pertussis is not well understood, as it is a severely underdiagnosed and underreported disease, with country surveillance data reported to the World Health Organization (WHO) totalling only 145,486 reported pertussis cases in 2019 [9].

Despite high infant vaccine coverage, pertussis is endemic in all countries and epidemic cycles have been reported every 2 to 5 years [2,10,11]. The regional distribution of pertussis cases exhibits significant variability in terms of incidence, mortality, and vaccination coverage, influenced by factors such as demographic profiles, geographic location, and vaccination policies. The seasonality of pertussis is unresolved, with evidence of a seasonal trend in some countries but none in others [12–14]. The transition from whole-cell pertussis vaccines (wP) to acellular pertussis vaccines (aP), which are less reactogenic but also less effective, has been linked to the changing epidemiology of pertussis. A shift in the age distribution of pertussis to older children, adolescents, and adults has been noted in some countries where aP vaccines have replaced wP vaccines [15]. The reported age shift may also be due to an improved awareness of less typical disease manifestations in older individuals, more sensitive and available laboratory testing, and better surveillance across all age groups [2]. Although pertussis is under-recognised in adults, there is increasing acknowledgement of its role as an important cause of disease in adolescents and adults with waning immunity.

Figure 1 shows the number of pertussis cases reported to the National Notifiable Diseases Surveillance System (NNDSS) in the United States of America (USA) from 1922 to 2023 [16]. A substantial decline is visible following the introduction of the diphtheria-tetanus-wP (DTwP) vaccine. However, an increase in cases can be seen following the introduction of aP vaccines. Although this shows the case pattern over time in the USA, similar patterns have been reported from other countries with extensive historic surveillance and shifts in vaccine formulation [17–19].

**Figure 1.**
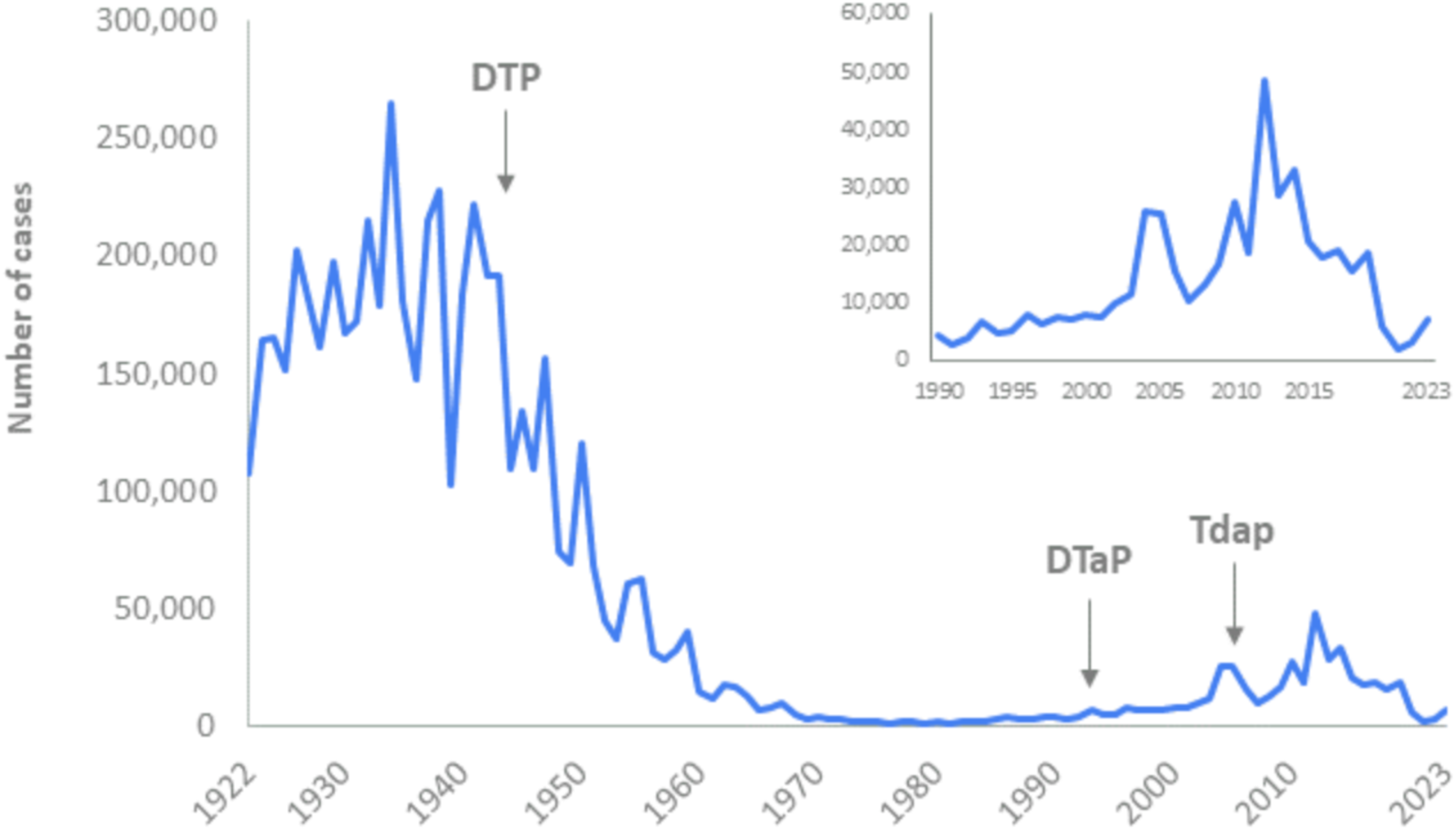
Reported NNDSS pertussis cases, United States of America, 1922–2023 [16] Data source: CDC, National Notifiable Diseases Surveillance System (NNDSS). DTP, diphtheria-tetanus-whole-cell pertussis vaccine; DTaP, diphtheria-tetanus-cellular pertussis vaccine; Tdap, tetanus-reduced diphtheria-acellular pertussis vaccine.

The resurgence of pertussis in highly immunised populations, particularly in adolescents and adults, is attributed to factors such as waning immunity, lack of booster immunisations, and improved diagnostic methods [4,20,21]. The Global Pertussis Initiative highlights the need for improved surveillance and tailored immunisation strategies, including booster doses for adolescents and adults, to address the persistent burden of pertussis globally [6]. Overall, the resurgence of pertussis in various regions underscores the importance of maintaining high vaccination coverage, particularly among infants and pregnant women, and adapting vaccination strategies to address waning immunity in older populations [22,23].

### 1.3 Pertussis vaccination and immunity

Two types of vaccines are available for pertussis: the wP formulation, which contains a suspension of killed bacteria, and the aP formulation, which contains 1–5 components of *B. pertussis*. Currently the wP formulation is used mainly in low-income countries (Figure 2) [24]. Due to reactogenicity of wP formulations, many high-income and some middle-income countries changed to aP vaccines.

**Figure 2.**
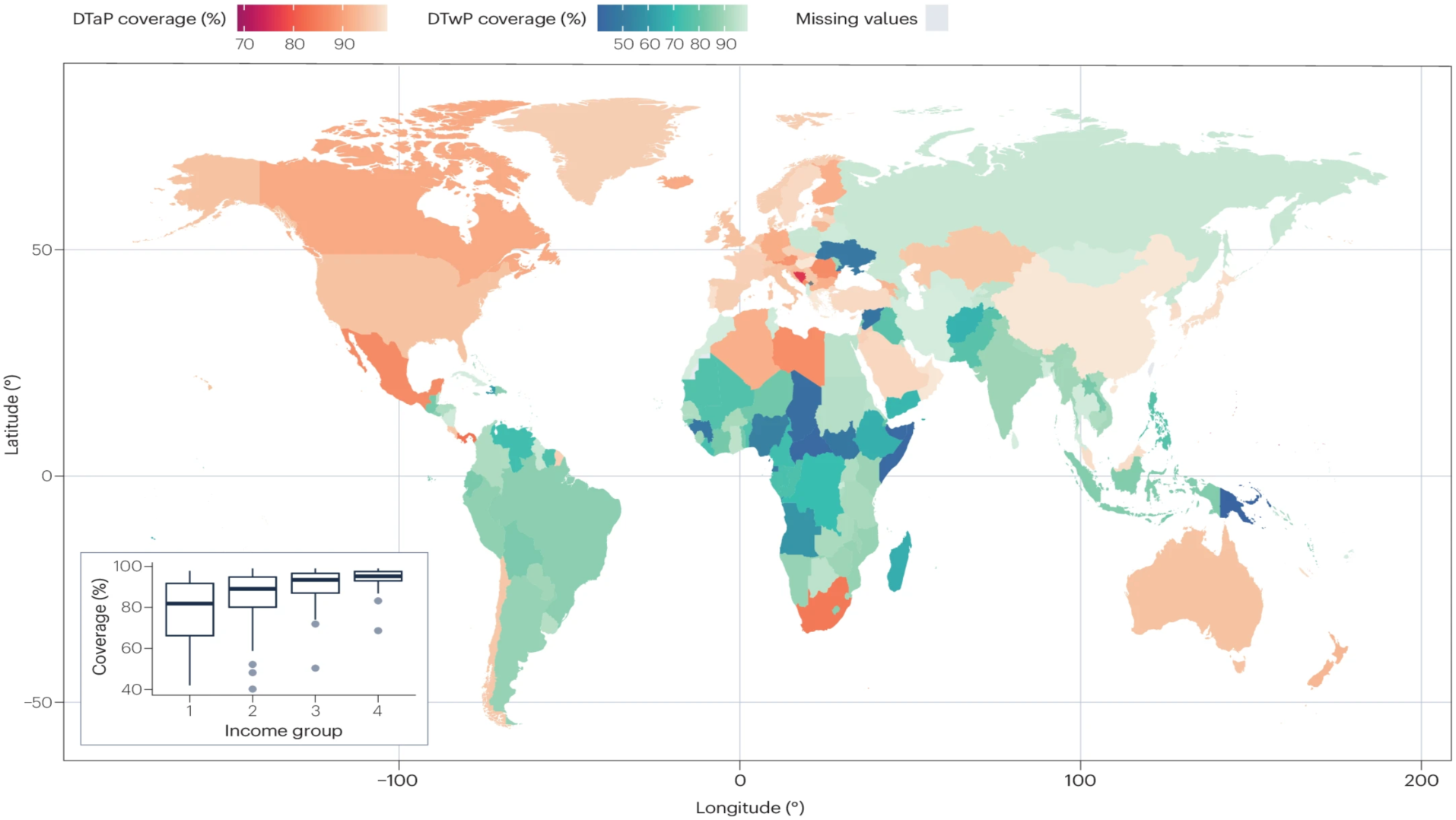
Map of average DTaP and DTwP coverage during 2015–2019 [24] Figure description: The different geographical areas are coloured according to the type of vaccine used for primary immunisation (red colours for DTaP vaccines; blue colours for DTwP vaccines). The inset represents vaccine coverage distribution in the different income groups. The figure is sourced from Domenech de Cellès and Rohani (2024), where the authors extracted data from WHO databases covering 208 geographical areas in the different income groups (low-income group, 28 areas; lower-middle-income group, 54 areas; upper-middle-income group, 54 areas; and high-income group, 72 areas).

There has been long-standing uncertainty around the duration and strength of immunity, both of vaccine-derived immunity and immunity following natural infection [14]. Neither infection nor vaccination confer lifelong or long-lasting protection; symptomatic reinfections can occur [2]. However, the precise nature of immune waning, immune failure, immune boosting (boosting the strength of immunity upon subsequent exposure to infection without experiencing a transmissible infection), and the variation of transmissibility between different types of infection is unclear [12,14,25–27]. There is also uncertainty regarding the importance of repeat infections in the transmission process and how response to pertussis exposure depends on different factors, for example age, time since last exposure, and vaccine composition [12]. These substantial gaps in the literature are compounded by challenges in surveillance, diagnostics, and reporting of pertussis [2]. Such challenges lead to severe under-reporting of pertussis and mean that the true infection and disease burden is not well understood.

Duration of immunity following a wP vaccine three-dose primary series schedule has been reported to range from 4 to 12 years, with some vaccines having a longer duration of protection [28]. Studies of aP vaccines report protection for 4 to 5 years following a three-dose primary series [28,29]. There is increasing evidence that protection following booster doses of aP vaccines wanes faster in individuals primed with aP compared to those primed with wP vaccines [28,30]. Epidemiological data show waning immunity in school-aged children, adolescents, and young adults in populations receiving an aP vaccine [31].

There is difficulty comparing estimates of efficacy and effectiveness of different vaccines given the significant heterogeneity across studies [2]. A systematic review calculated a pooled efficacy of wP vaccine against pertussis disease in children of 78%, with significant variation among vaccines [32]. Little is known about the effectiveness of wP vaccines in older age groups. Direct effectiveness data for vaccines that are currently in use are not available. Observational studies have consistently shown around 50% protection against *severe* pertussis in infancy following a single dose of either wP or aP vaccine, and that two doses offer at least 80% protection against severe disease [2]. There is mixed evidence of the relative efficacy of aP and wP vaccines [2]. Studies to date indicate that aP vaccines are more effective than low-efficacy wP vaccines (wP vaccines shown to be suboptimal are no longer in use), but may be less effective than the highest-efficacy wP vaccines [2]. Studies comparing wP and aP vaccines in non-human primate models [33,34] demonstrated that aP vaccines protect against severe pertussis-associated symptoms but not against infection or colonisation. In addition, the aP-vaccinated baboons did not clear infections quicker than unvaccinated animals and, because they failed to prevent colonisation, were able to transmit *B. pertussis* infection to contacts. In contrast, wP-vaccinated animals cleared infection more rapidly and generated a better T-cell response protecting better against colonisation and shedding [33,34].

Vaccinating mothers during pregnancy with combined tetanus, diphtheria, and acellular pertussis (Tdap) has proven highly effective in protecting newborns by transferring antibodies, significantly reducing laboratory-confirmed disease in unvaccinated infants [4,35–37]. Despite concerns about potential immunological blunting in infants, epidemiological evidence supports the effectiveness of maternal immunisation. Vaccination of pregnant women has been implemented with the existing aP-containing vaccines for adult use in several industrialised countries with an impressive impact on infant disease [35–37]. Several studies confirmed the benefit of vaccination during pregnancy with documented high antibody concentration in neonates persisting until primary vaccination is started [38–41]. Vaccination should ideally occur at least two weeks before delivery. Data from Switzerland indicated that immunisation in the second trimester is associated with higher levels of pertussis antibodies in cord blood compared to third trimester vaccination [42]. In the UK, the vaccine effectiveness was 95% against pertussis-associated deaths and ∼90% against hospitalisations in infants <3 months old, with a stronger effect when vaccination was performed with larger intervals before delivery [35–37]. Similar results have been reported from several other countries where this strategy has been implemented [23,43–45].

Seroprevalence studies can be valuable tools for understanding the true burden of infectious diseases. However, for diseases without a clear serological correlate of protection and with complex immune dynamics, the interpretation of seroprevalence studies can be challenging. This is the case for pertussis, which exhibits the phenomenon of immune boosting—where exposure to the pathogen can trigger a detectable immune response without causing a transmissible infection. Unlike diseases with clear antibody thresholds for immunity (e.g. 0.1 IU/mL for diphtheria toxin), pertussis lacks a well-defined protective titre [46]. Immunoglobulin G (IgG) antibodies against pertussis toxin (PT)—a key virulence factor of *B. pertussis*—are measured as a surrogate of pertussis immunity. Serological assays report anti-PT IgG, often using thresholds to categorise exposure or immunity. For example, seropositivity cut-offs ranging from ≈5 IU/mL (near the assay detection limit) up to ≈50 IU/mL have been used to indicate the presence of pertussis antibodies (and thus some degree of immunity) [46]. Higher antibody cut-offs, usually ≥100 IU/mL, are commonly interpreted as evidence of recent pertussis infection or exposure within roughly the past year [46,47]. Despite these conventions, antibody levels alone do not guarantee protection, and low titres do not necessarily equal susceptibility. Protection is measured probabilistically—higher anti-PT IgG generally correlates with lower risk of disease, but there is no absolute’immune/not immune‘cutoff. A study in Sweden demonstrated that children with higher post-vaccination PT IgG were far less likely to develop pertussis: among children exposed to pertussis at home, those who remained healthy had a median anti-PT IgG of 246 IU/mL, versus 79 IU/mL in those who fell ill with severe pertussis [48]. This three-fold difference in antibody levels was highly significant, underscoring that higher pertussis antibody titres correlate with protection. However, because pertussis immunity also involves cellular immunity (e.g. T-cell memory), serology captures only one aspect of protection [49]. In practice, protection is usually defined where the presence of anti-pertussis antibodies (especially anti-PT IgG) is taken as a marker of immunity, and very high titres signal recent boosting of immunity, but no firm protective threshold has been universally adopted.

Resolving the issues surrounding pertussis burden is challenging due to atypical presentation, reliance on clinical suspicion, the absence of a serological marker for immunity, limited bacterial genome sequencing, and heterogeneous diagnostic methods. Further complicating matters, studies suggest that asymptomatic infections, though detectable, have unclear implications for onward transmission [24]. On the one hand, asymptomatic carriers can transmit pertussis (particularly within families), although at lower rates than fully symptomatic cases, thereby contributing to the circulation of pertussis even in highly vaccinated populations. On the other hand, asymptomatic exposures serve to boost immunity in the population, either with or without a transmissible infection. This can help reduce the probability of subsequent symptomatic infection but also complicates the interpretation of seroprevalence data.

### 1.4 Mathematical models of pertussis

There is an extensive body of literature using mathematical models to examine the mechanisms of pertussis epidemiology and explore the likely impact of different vaccination strategies. Given the high degree of uncertainty regarding key features of pertussis biology, immunology, and incidence, there are many different model structures, and varying approaches to calibration across these studies. Campbell et al. provide a comprehensive review of pertussis models representing the breadth of key model features in the literature, critically discussing the approaches adopted as well as the impact of the selected model structures and assumptions on the study conclusions [50].

The rate at which the susceptible population acquire infection (force of infection, λ) depends on contact frequency and the likelihood of transmission. Early models used a single value for transmission rates, while more recent models break it down into constituent parts [12,51,52]. Regarding infectiousness, the simplest assumption is that all infections contribute equally to the force of infection [53]. However, more complex models allow for varying levels of infectiousness based on immune state, infection type, or even non-infectious boosting cycles [54–57]. Models address how prior infection and vaccination modify infection and immunity [55,57–59]. Assumptions range from individuals being fully protected or fully susceptible [53,60], to partial protection where susceptibility is reduced [54], or enhanced boosting where partially waned individuals are more likely to mount an immune response [56,57].

It is common for pertussis models to stratify compartments into discrete age groups to model vaccination schedules or age-specific differences in behaviour and infection characteristics. Contact matrices characterise transmission between subgroups, either explicitly or implicitly. Homogeneous mixing assumes the susceptible population acquire infection from all infected individuals at the same rate [53], while age-structured models can account for different transmission rates within and between age groups [61]. Empirical contact data is increasingly used to inform these matrices.

Pertussis exhibits epidemic cycles with periodicity typically ranging from two to five years. These recurrent outbreaks are driven by a complex interplay between host immunity, pathogen transmission dynamics, and demographic processes. Following natural infection or vaccination, immunity against pertussis wanes relatively rapidly, often within 4–12 years, resulting in a growing pool of susceptible individuals in the population [27,57]. This accumulation of susceptibles eventually surpasses the threshold necessary for sustained transmission, triggering epidemic waves. Birth rates play a central role in replenishing the susceptible population, especially among infants too young to be fully immunised. In addition, heterogeneity in vaccine-induced versus natural immunity contributes to variations in protection and transmission potential [52]. Mathematical modelling and epidemiological data suggest that these dynamics resemble the cyclical patterns observed in other childhood infections such as measles, but with more irregular periodicity due to the shorter duration of pertussis immunity and variable vaccine coverage [62]. Thus, pertussis epidemic cycles emerge from the interaction of waning immunity, demographic turnover, and stochastic fluctuations in transmission, producing characteristic outbreaks every few years despite ongoing vaccination programmes. Some models generate these ongoing cycles as a natural consequence of transmission dynamics without explicit seasonal forcing or stochasticity [63,64]. Other models incorporate’seasonal forcing’ (seasonal variation in transmissibility) or stochasticity to generate these cycles [51]. Table A.1 (see Appendices) provides a summary of the pertussis models and key features.

## 2. Materials and methods

This section presents an overview of the age-structured, compartmental pertussis model, equations, key assumptions, and parameters. Model development and parameters were informed by engagement with a technical expert group (see Appendices). Figure 3 shows the conceptual model diagram for pertussis with a list of corresponding model compartments. Full compartment descriptions are provided in Table A.2. Tables 1 and A.3 detail the model parameters for pertussis.

**Figure 3.**
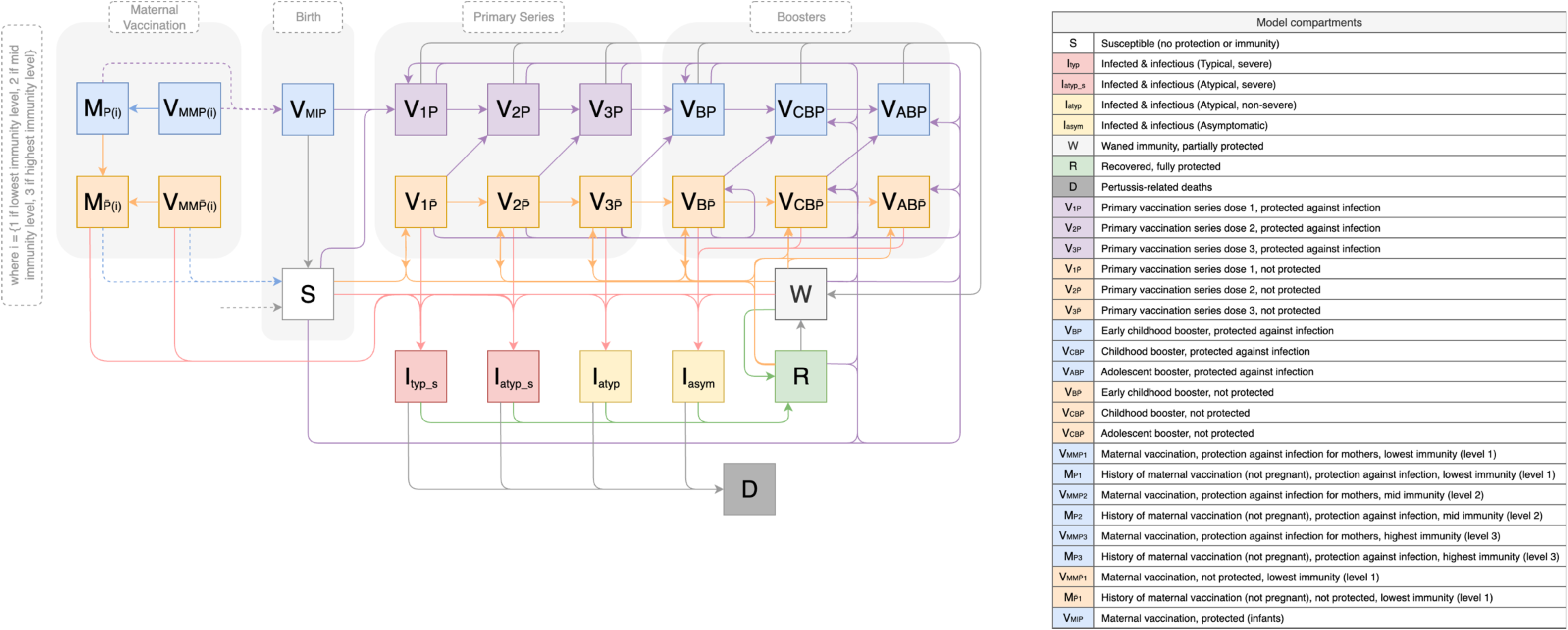
Conceptual diagram for pertussis model structure

### 2.1 Model structure

The model is a compartmental transmission model with 29 states, stratified by 56 age groups (*i*). Age categories were chosen to simulate vaccination and immunity at key ages; from very fine age groups (several weeks or months during the first two years of life), to annual age groups through to 15-years old, then five-year age bands until >75-years old. For simplicity, age subscripts are not included in the model description below but are shown in the model equations for clarity. The full list of model age groups is in the Appendices (Table A.2).

Fully or partially susceptible population (S, W, and any non-protected vaccination state) of all ages can acquire infection through contact with an infectious person and move to one of the infected states (I_0_ = typical, severe infection, I_1_ = atypical, severe infection, I_2_ = atypical, non-severe infection, I_3_ = asymptomatic infection) depending on age. The infectious states (I_0_, I_1_, I_2_, I_3_) capture distinct types of infection that have different mortality, infectiousness, and symptoms (see Table A.3 for details). Populations in these states are infected and infectious. They remain in the state for the duration of infectiousness.

Upon recovery from an infected state, the recovered population moves to R (recovered, fully immune). As naturally acquired immunity wanes, previously protected populations transition to W, which is a waned state of partially protective immunity. Upon exposure to an infected individual, those in W can either become infected (moving to I_0_, I_1_, I_2_, I_3_) or experience immune boosting, enabling a move from W directly into R without experiencing a transmissible infection. This boosting effect is in line with Lavine et al.’s work on modelling pertussis immunity [57]. The proportional split between I_0_, I_1_, I_2_, I_3_ and boosting to R is determined by age and type of vaccination, where aP allows a higher probability of symptomatic infection than wP or natural infection.

The vaccine states in the model cover maternal immunisation (V_mm_), the diphtheria, tetanus, pertussis-containing vaccine (DTPCV) primary series (V_1_, V_2_, V_3_), and the three WHO-recommended DTPCV booster doses (V_b_, V_cb_, V_ab_). For all vaccinated states, the protected/not-protected split is based on the effectiveness (ε) of each dose of the primary series or boosters for protection against infection. Individuals are born into the model into one of two states: V_MI_p_ or S. The proportion (based on vaccine coverage) of infants born to mothers who receive maternal vaccination against pertussis are split by the vaccine effectiveness (ε) into V_MI_p_ (mother receives vaccine(s) during pregnancy and the infant is protected) and (1-ε) into S (mother receives vaccine(s) during pregnancy but the infant is not protected). As we assume that there is no immune blunting from maternal vaccination, infants born to non-vaccinated mothers and those born to vaccinated mothers but not protected, both enter straight into S.

Upon vaccination with the first dose of the primary series, the vaccinated population moves into V_1_p_ or V_1_np_ depending on vaccine effectiveness, and so on for future doses. we assume that the primary series is cumulative, such that the eligible population would only receive higher doses in the schedule if they have received the previous doses (i.e. can only move into V_2_ if moving from V_1_ not S). We assume that the booster doses are not cumulative, such that the appropriate population can receive a booster dose when eligible by aging irrespective of the compartment of origin. For the primary series and boosters, we assume that those moving from a protected vaccination state to a higher dose state will remain protected by the vaccination (i.e. V_1_p_ will only ever move into V_2_p_, not V_2_np_). However, those moving from a non-protected state may move into the protected or non-protected state of the subsequent dose, dependent on vaccine effectiveness. Over time, vaccine-induced immunity wanes allowing this population to transition to W. The level of protection in W is the same for those waning from naturally acquired and vaccine-induced immunity. Those in the non-protective vaccine states (V_J_np_) will remain in that state unless a transition is triggered by infection or vaccination.

Maternal vaccination is modelled to be offered to pregnant women based on age-specific fertility rates via antenatal care visits. Protection following maternal vaccination is modelled through eight states for mothers and one state for infants, who are protected by maternal antibodies after birth. The eight states for mothers are stratified into three levels of protection: lowest immunity (level 1), mid immunity (level 2) and highest immunity (level 3); with the states capturing whether the mother has been vaccinated for the current pregnancy or a previous pregnancy, and whether this provided protection, based on vaccine effectiveness. The immunity level (1 to 3) following maternal vaccination then depends on the prior state of immunity (history of vaccination with the primary series and booster doses). This allows the model to retain the level of immunity and corresponding duration of protection. After the nine-month period of pregnancy, there is a move from V_MMP_*_k_* and V_MM<u>P</u>_*_k_* into M_P_*_k_* and M<u>_P_</u>*_k_*, respectively. This captures the history of maternal vaccination and allows women to retain vaccine-derived protection beyond the period of pregnancy. Individuals move from a protected state (M_P_*_k_*) into an unprotected state (M<u>_P_</u>*_k_*) based on the waning rate of the corresponding immunity level.

Pertussis incidence in many countries shows epidemics cycles of 2–5 years. To capture this oscillation, in keeping with Hethcote’s approach [54,55], we incorporate periodicity and stochasticity into the model, which is calibrated to the local setting patterns (Equation A.31).

##### Box 1. Summary of key model assumptions for the pertussis model

For infection and transmission, the following assumptions are made:

- Pertussis is transmitted person to person, through respiratory droplets and close physical contact.
- There are four types of infection, which differ by presentation, infectiousness, and severity.
- Probability of infection type depends on age, immunity level, and type of vaccination received (wP vs aP).
- Both symptomatic and asymptomatic individuals can transmit pertussis but with differing levels of infectiousness.
- Asymptomatic individuals have a lower relative infectiousness than symptomatic individuals.

For vaccination and immunity, the following assumptions are made:

- Immunity can be obtained from previous infection, vaccination or both.
- Both vaccine-derived and naturally acquired immunity are assumed to wane over time.
- Whole-cell vaccines are more effective than acellular vaccines in protecting against transmission and disease. In particular, wP provides greater, but not full, protection against both transmission and disease than aP.
- Upon exposure to an infected individual, those with partial immunity (W) can either become infected or experience immune boosting (moving directly to a recovered, fully protected state without experiencing a transmissible infection).
- There is no immune blunting from maternal vaccination.

### 2.2 Model parameters

Table 1 shows the description and dimensions (time, age, and infection type) of the model parameters for pertussis. Table A.4 (Appendices) provides the unit, range, distribution, and source of the parameter values for pertussis. Parameter values were informed by literature and discussions with disease experts who formed part of a technical expert group (TEG).

**Table 1.**
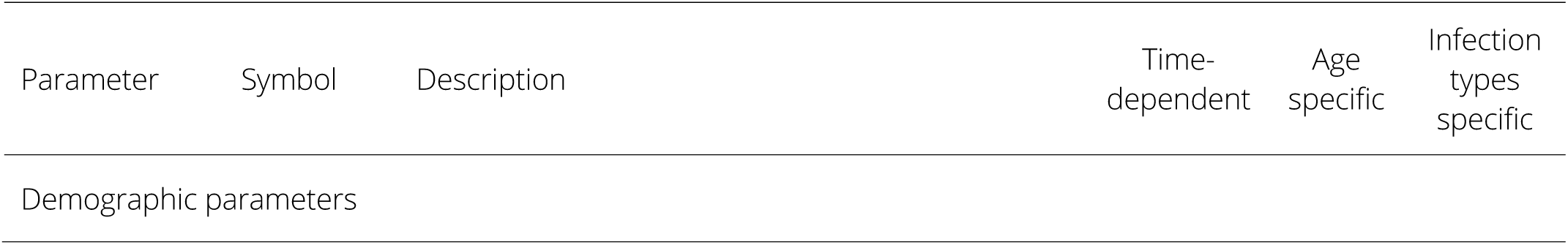
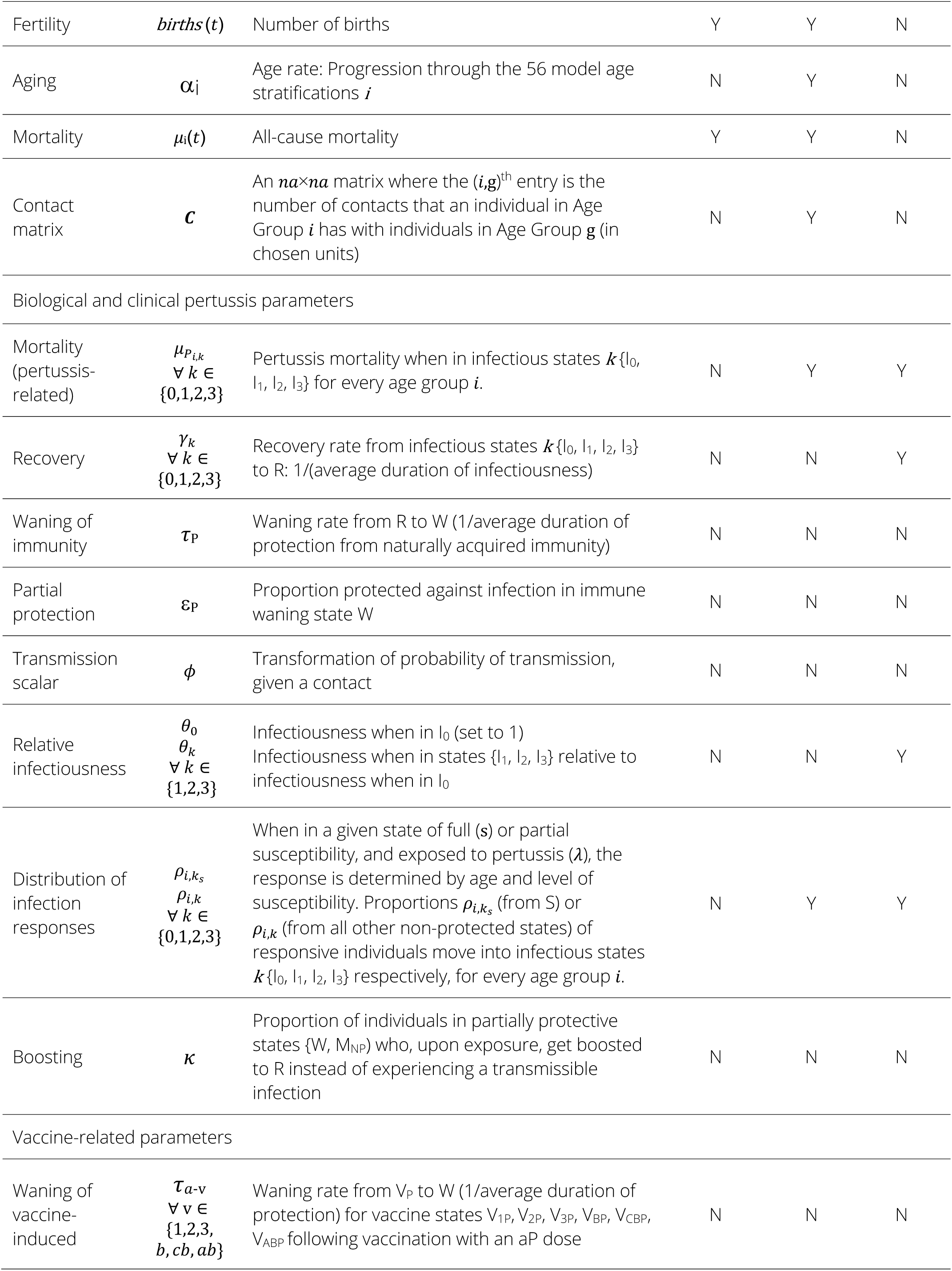
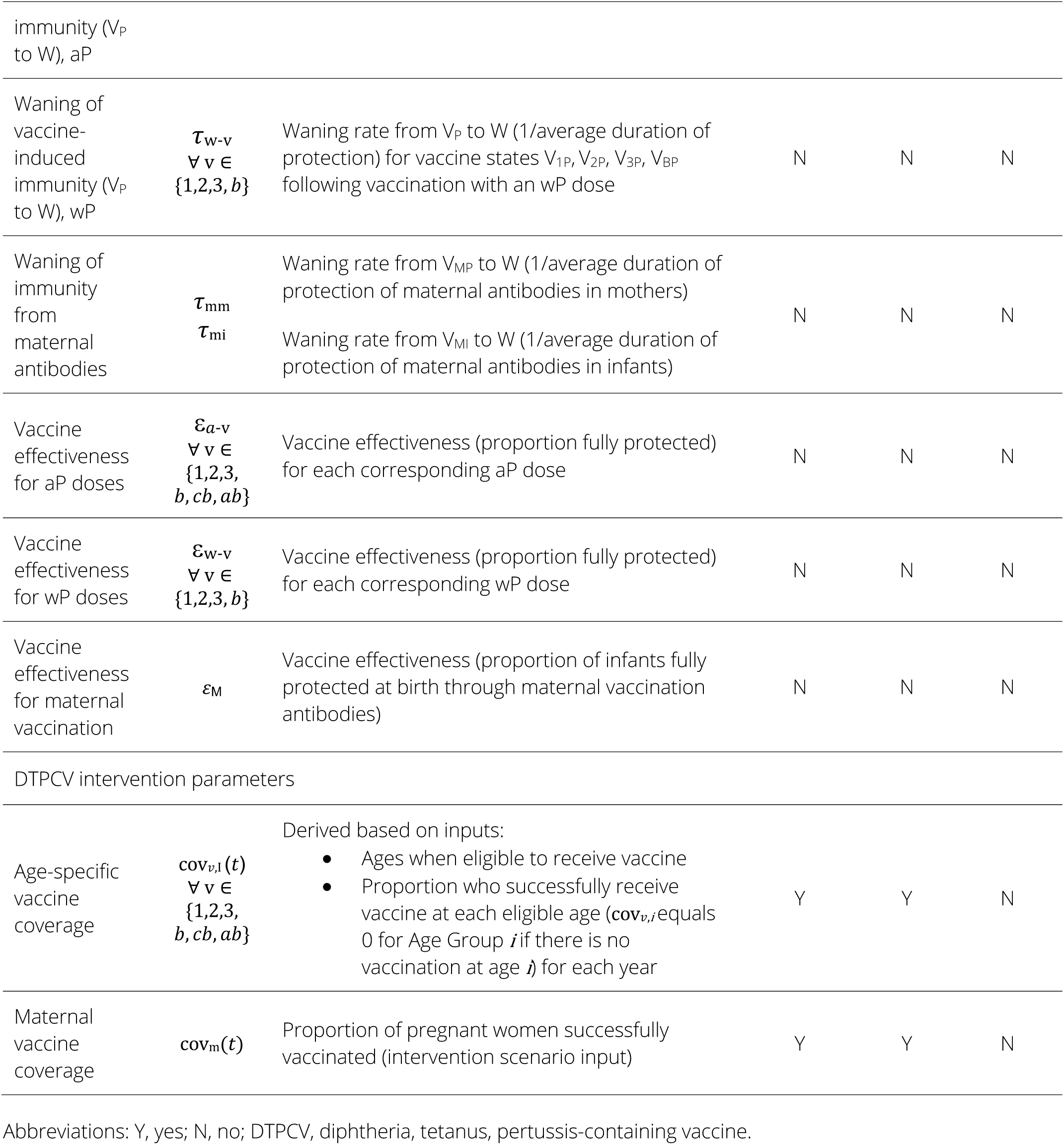
Description and dimensions of model parameters for pertussis.

### 2.3 Sensitivity analysis

We conducted a sensitivity analysis (SA) on the pertussis model using Latin hypercube sampling (LHS)-partial rank correlation coefficient (PRCC) [65–67]. Drawing from the appropriate parameter distributions (Table A.4), we conducted LHS of the parameter space to generate 1000 parameter sets. Input variables for the PRCC are pertussis transmission scalar; likelihood of death; relative infectiousness of infection types I_1_, I_2_, and I_3_ compared to I_0_; recovery rate of infection types I_0_, I_1_, I_2_, and I_3_; boosting proportion; immunity waning rate (infection); immunity waning rate (vaccine); waned protection; and vaccine effectiveness. Three model outputs (average annual pertussis incidence, proportion of the population protected against pertussis, and average annual number of pertussis deaths) for three age groups (all ages, under 5-year-olds, and under 15-year-olds) are considered for the PRCC.

## 3. Results

### 3.1 Sensitivity analysis findings

Modelled incidence and deaths are most sensitive to changes in immunity waning rates for naturally acquired and vaccine-derived immunity, the boosting proportion, and the recovery rate of severe atypical infections (I_1_). This is consistent across the three age categories considered. Modelled estimates of population protected are most sensitive to changes in the transmission scalar, immune boosting parameters (boosting proportion and waned protection), and the relative infectiousness of asymptomatic infections, across the three age categories. The sensitivity of population protection to the immune waning rate for vaccine-derived and naturally acquired protection varies by age category. The immunity waning rate of vaccine-derived protection is the third most sensitive parameter in under 5-year-olds, the seventh most sensitive parameter in all ages, and is not significant in the under 15-year-olds. The full sensitivity analysis is presented in the Appendices.

### 3.2 Key model features and dynamics

This section explores key transmission and immunity dynamics in the model. Section 3.2.1 demonstrates the role of immune boosting in the model, investigating the interplay between the two immune boosting parameters. Section 3.2.2 explores immune waning, showing the breakdown of vaccine-derived and naturally acquired immunity in the model. Section 3.2.3 presents the clinical profile of infection by age.

#### 3.2.1 Immune boosting

Figure 4 shows the variation in the proportion of the population protected against pertussis with different values for the two key boosting parameters: waned protection and boosting proportion. Waned protection (λ_W_) captures the proportion of the population in the partially protected/waned states who are protected from infection. The boosting proportion (Κ) captures the proportion of the population in partially protective/waned states who, upon exposure, get boosted to R instead of experiencing a transmissible infection.

**Figure 4.**
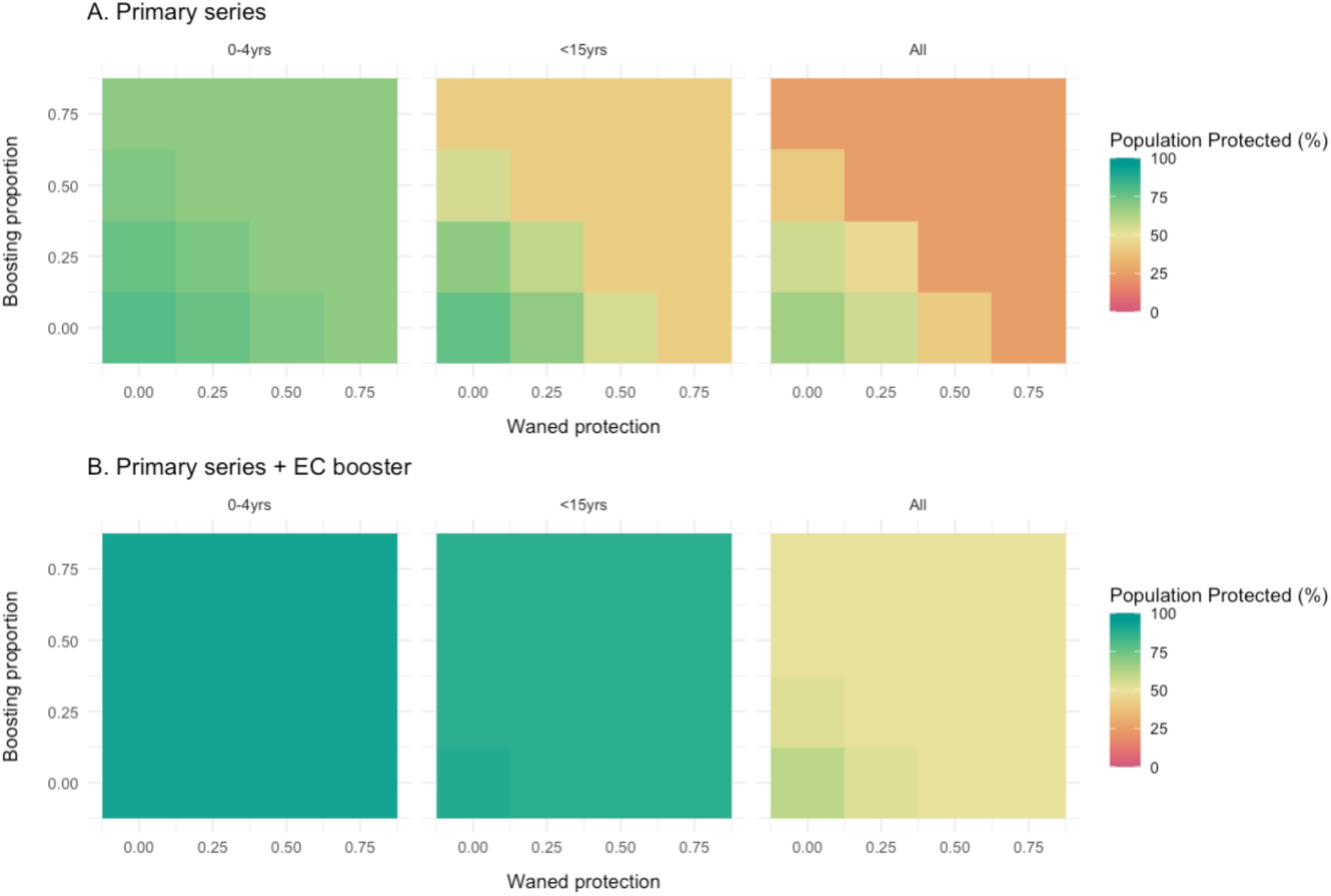
Sensitivity analysis of pertussis immunity to changes in immune boosting for the DTP primary series and EC booster scenarios Figure description: Full pertussis immunity is defined as the proportion of the population in either protected vaccination states or the recovered state. The impact of immune boosting is explored where 1) the boosting proportion is the proportion of the population who, when exposed to an infectious contact, get boosted to R without experiencing a transmissible infection, and 2) waned protection is the proportion of the population in waned compartment W who are protected from infection. The top panel shows a scenario with the primary series only and the bottom panel shows a scenario with the primary series plus an early childhood booster dose. EC, Early childhood booster; R, Recovered model compartment; W, Waned model compartment.

We ran the model 16 times with different parameter combinations for λ_W_ and Κ (from 0 – 0.75), for a scenario with the primary series (top panel A) and the primary series plus the early childhood booster (bottom panel B). Figure 4 demonstrates that immune boosting has a more sizeable effect in the scenario where only the primary series is given, and that the impact in this scenario is more pronounced in the total population than younger age groups. This follows expectations, as vaccination with the primary series provides protection in younger ages, leaving less opportunity for impact on immunity via boosting, which only acts on the waned proportion of the population in the model. This is even more pronounced with the introduction of the EC booster, which extends the duration of vaccine-derived protection.

Figure 5 shows the results from the same set of runs as Figure 4 but considers the impact of boosting on clinical incidence per 100,000 population for the DTP primary series only. The pattern is the same as in Figure 4, but the effect is more pronounced. In the absence of boosting, where both the boosting proportion and waned protection are zero, the highest clinical incidence is projected. The impact is greatest in younger age groups, as these are the ones more susceptible to clinical infection (adults are more likely to experience an asymptomatic infection than children). Beyond a certain threshold, boosting has no further effect on population protected, as can be seen in the upper right diagonal of the panels in Figures 4 and 5.

**Figure 5.**
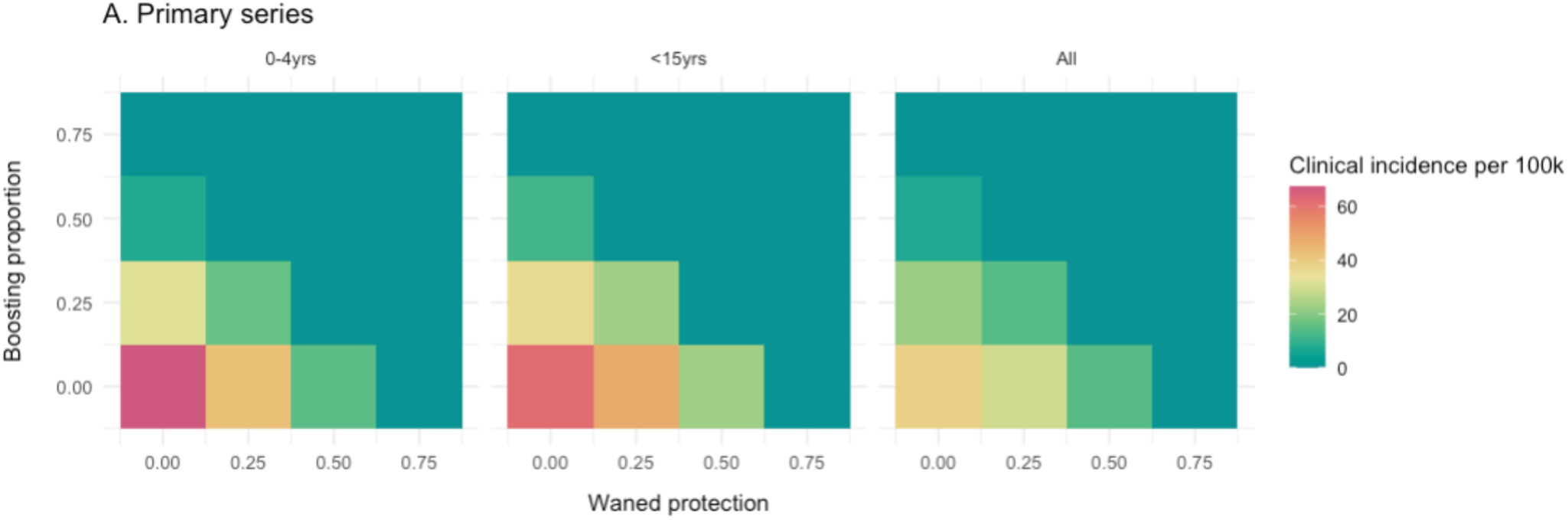
Sensitivity analysis of projected clinical incidence of pertussis per 100k population to changes in immune boosting for a scenario with the DTP primary series only Figure description: Clinical incidence per 100k includes all symptomatic cases projected by the model (i.e. excluding the asymptomatic compartment, I_asym_). The impact of immune boosting is explored where 1) the boosting proportion is the proportion of the population who, when exposed to an infectious contact, get boosted to R without experiencing a transmissible infection, and 2) waned protection is the proportion of the population in waned compartment W who are protected from infection. The model is run for a scenario with the primary series only. R, Recovered model compartment; W, Waned model compartment.

#### 3.2.2 Immune waning

Figure 6 shows a breakdown of vaccine-derived and naturally acquired immunity against pertussis in a hypothetical setting with the primary series and no DPTCV booster doses. To investigate vaccine-derived immunity, we simulated an artificial setting with no pertussis transmission (only vaccination) and compared this to a setting with transmission and vaccination. The model was run on the same parameter set in both scenarios, with the only difference being the presence (light blue) or absence (green) of infection. Over time, both naturally acquired and vaccine-derived protection wane. Vaccine-derived immunity shows the expected decline in protection as protection from the primary series wanes. An increase in vaccine-derived immunity is visible in the reproductive ages due to the presence of maternal immunisation. Naturally acquired immunity and immune boosting play an important role in the total level of protection. This is especially prominent in older age groups, when vaccine-derived immunity has waned and most protection is from infection or boosting. In practice, it is extremely challenging to differentiate the source or nature of protection [24].

**Figure 6.**
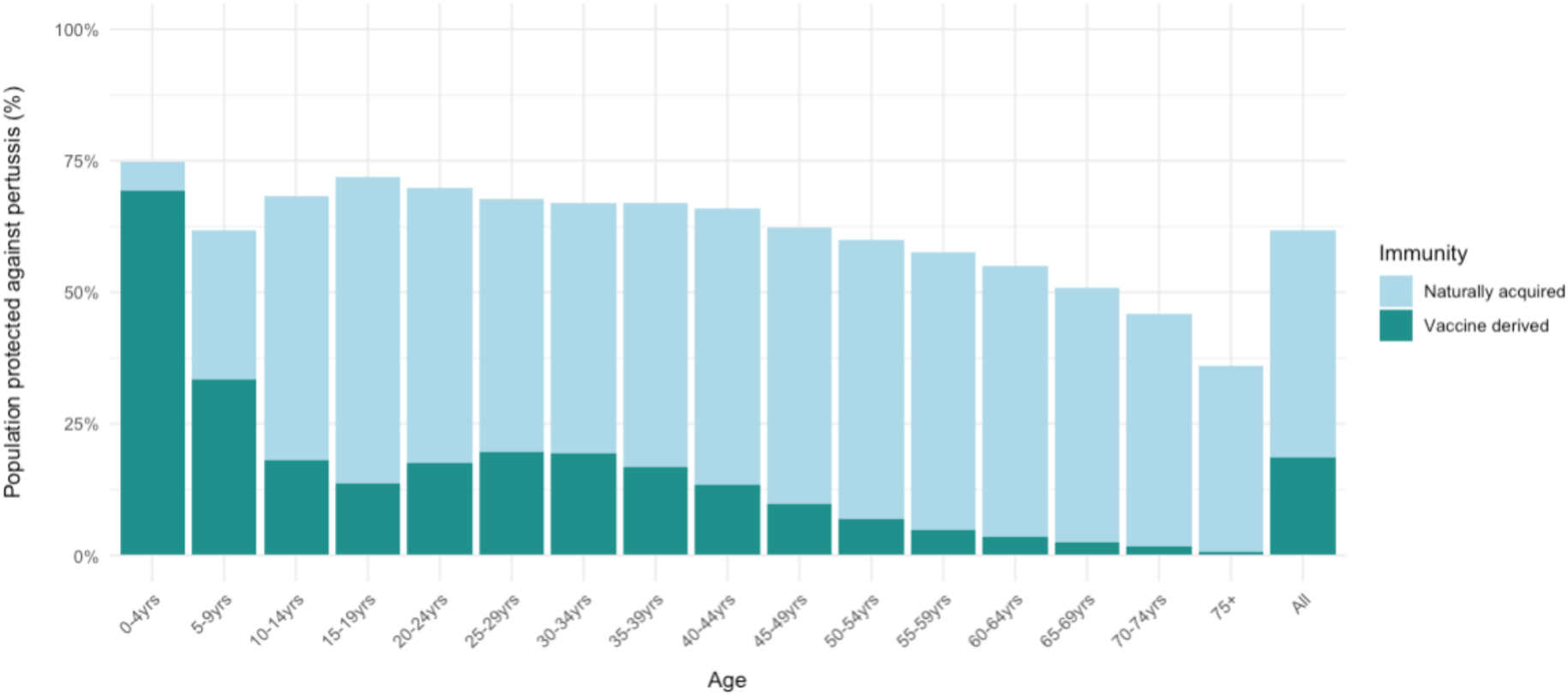
Comparison of vaccine-derived and naturally acquired immunity against pertussis

#### 3.3.3 Clinical profile

Figure 7 shows the breakdown of clinical presentation (asymptomatic, mild, and severe) of pertussis incidence by age, per 100,000 population. Generally, younger children are more at risk of infection and disease and tend to experience more severe presentations of the disease than older children, adolescents, and adults. This is captured in the model through an age-stratified probability of infection type (Appendix 4A). If the study setting has reliable case data or sentinel surveillance site data, the model can be fitted to replicate the exact age distribution of cases. In the absence of this, the general pattern below replicates what is described in literature and from discussions with disease experts.

**Figure 7.**
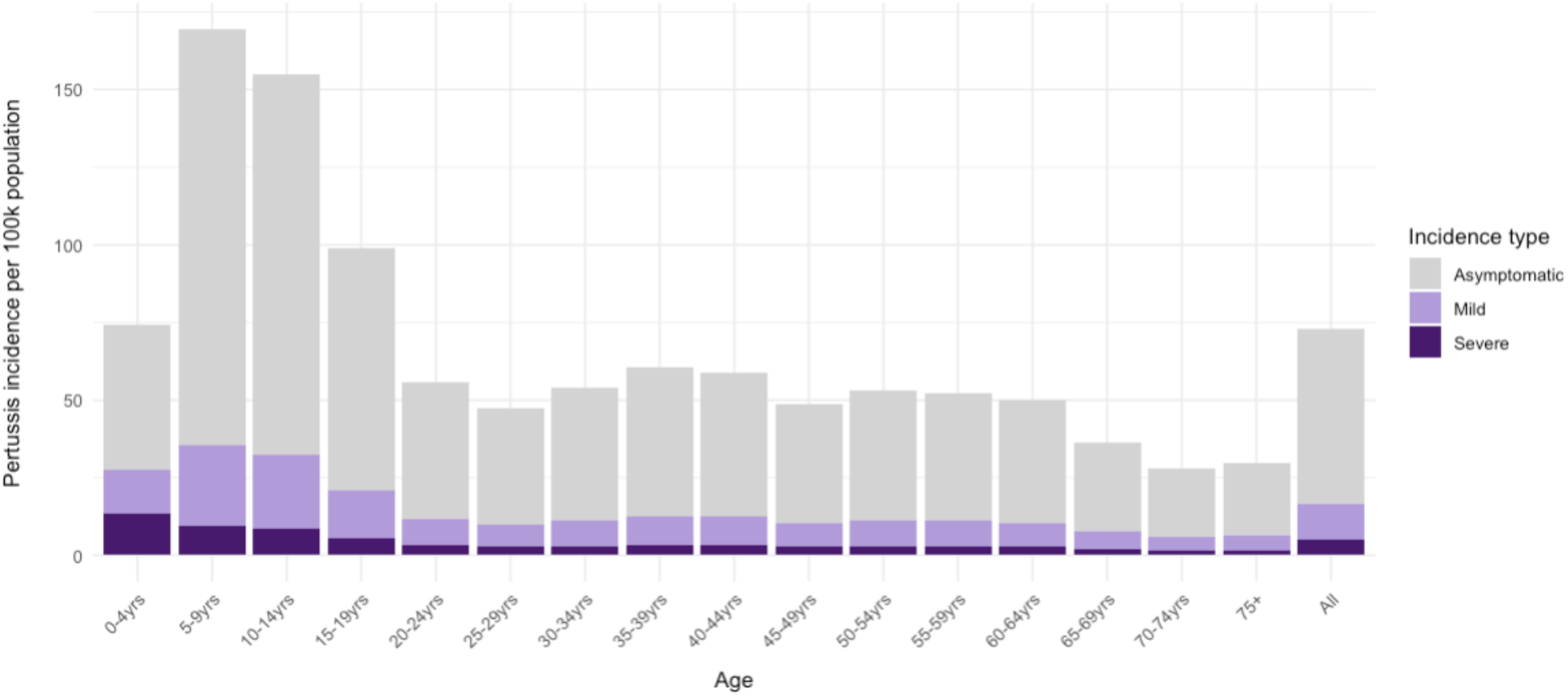
Asymptomatic, mild and severe pertussis incidence per 100,000 population, by age Figure description: The bars show model projections of pertussis incidence per 100,000 population, in 5-year age bands. The incidence categories map to the four infection compartments in the model as follows: Asymptomatic = I_3_ (I_a_), Mild = I2 (I_atyp_ns_), Severe = I_1_ (I_atyp_s_) + I_0_ (I_typ_).

## 4. Discussion

### 4.1 Key finding and features

We have developed an age-structured, deterministic compartmental model of pertussis and explored its key dynamics. A review of pertussis models to inform policy highlighted several features that are critical for models to include to capture key dynamics of pertussis [50]. These are age structure, waning immunity for vaccine-derived and naturally acquired protection, reinfection of previously immune population, immune boosting, and age distribution of prior vaccination schedules and coverages. We have addressed each of these key features.

The model is stratified into 56 age groups, allowing detailed vaccination schedules, age-dependent parametrisation, and model outputs by age. This is important for pertussis, where severity is greatest in infants and clinical presentation changes with age [2,4,11,24]. The model also includes four different infection types (severe typical, severe atypical, non-severe atypical, asymptomatic), which capture variations in the severity, presentation, and level of infectiousness. This represents the highly heterogeneous nature of clinical presentation of pertussis and enables the model to explore the role of asymptomatic infections in transmission and immunity patterns. Table 1 shows the parameters that are time-dependent, age-dependent, and specific to each of the four modelled infection types. The age-dependent parameters in the model are fertility, mortality (all cause and pertussis-related), contact mixing patterns, and distribution of infection types. While fertility and mortality data are available with detailed age-stratification by country, there are limitations in the available data required to adjust contact matrices by age and parametrise the age-breakdown of the distribution of infection types. Nevertheless, key patterns can still be incorporated. For example, the following principles are captured in the model’s distribution of infection type: 1) individuals with some immunity (vaccine-derived or natural) will experience less severe infection; 2) as age increases, severity of disease decreases; 3) the type of severe infection (typical or atypical) changes with age—up until 6-months old, in the absence of vaccination, there is a higher likelihood of experiencing an atypical severe infection, after which there is a higher likelihood of experiencing a typical severe infection; and 4) there will be a smoother transition of values for unprotected than for vaccinated as the protected will receive jumps in immunity in line with vaccination. In the absence of specific country data, the model replicates these main principles [2,11,24].

### 4.2 Contributions

The model captures several important immunity-related dynamics, including dose-specific waning of vaccine-derived protection; waning of naturally acquired protection; a state of waned partial protection, which allows reinfection of the population that has prior immunity; and immune boosting, which allows an increase in immunity without experiencing a transmissible infection. While the exact duration of vaccine-derived and naturally acquired infection, especially for booster doses, is not conclusively established in the literature, there is consensus that neither type of immunity is lifelong [2,14]. In line with other advanced, policy-focused models of pertussis, we use exponential waning to model the decline in protection [12,68]. Unlike simpler models that model protection in a binary manner, we include a waned compartment that offers partial protection. The strength of this partial protection is an important driver of incidence and protection in our model.

The model explicitly includes each dose of the primary series; booster doses in early childhood, childhood and adolescence; and maternal vaccination. It captures detailed historical vaccination schedules and coverage, allowing for differences in model behaviour for aP versus wP formulations, and changes between the formulations over time. This enables previous cohort effects to inform the current profile of protection and transmission.

Campbell et al. argue for the use of multivariate sensitivity analysis, noting that many published models of pertussis often vary only one or a few parameters at a time [50]. Our sensitivity analysis employed LHS-PRCC, allowing variation of a large set of parameters over plausible ranges. We found that incidence and deaths in the model are most sensitive to changes in immunity waning rates for naturally acquired and vaccine-derived immunity, the boosting proportion, and the recovery rate of severe atypical infections.

### 4.3 Limitations

Although we ran a comprehensive multivariate sensitivity analysis, we did not explore the impact of changing assumptions for the contact matrix or the distribution of infection type. For the contact matrix, this could have an impact on the model’s projections for infection and mortality in infants, who are most susceptible to severe presentations of pertussis. The contact patterns for infants (<2-year-olds) are likely to be different to those in early childhood (2–5-year-olds), given patterns in childcare, social interactions, and pre-school attendance. However, the contact matrix used is presented in five-year bands [69,70], which we modify based on country-specific demographics and the age-stratification in the model. While this is a reasonable assumption in the absence of detailed empirical data, it may not be a true reflection of reality. Several studies have highlighted the importance of contact mixing assumptions [50,61,71], so this would be an area for future investigation in the model.

There is substantial uncertainty in the literature surrounding the role of asymptomatic infections in onward transmission. While their prevalence is acknowledged, their precise contribution to overall transmission dynamics remains a significant point of contention in pertussis epidemiology [24]. Asymptomatic infections represent a potentially large but poorly characterised mass of infections that are purported to explain pertussis epidemiology [24]. Van Boven et al. modelled waning immunity and sub-clinical infection to explore the implications for pertussis in The Netherlands [60]. Their analysis indicated that the contribution of sub-clinical infection in adults to the circulation *B. pertussis* in The Netherlands could only have been significant if the vast majority of adult infections were sub-clinical [60]. Broadly, there are two main views regarding the role of asymptomatic infections. One posits that pertussis vaccines, particularly DTaP, offer only short-term protection against infection, leading to a large pool of asymptomatically infected adults who then transmit to susceptible children. The other estimates long-lasting effectiveness of vaccines, including DTaP, with only a modest transmission contribution from adults. Our model explores the sensitivity of asymptomatic contribution and waning immunity. It is robust to changes in the relative infectiousness of asymptomatic infection for incidence and deaths (Figure 2). However, the proportion of population protected is sensitive to changes in the assumption of relative infectiousness of asymptomatic infections. The parametrisation of the distribution of infection type was informed through extensive discussion with the TEG. While this provided consensus and a degree of confidence in the values used, conducting a sensitivity analysis on different assumptions for the distribution of responses would offer a more comprehensive view of the role of asymptomatic infections in the model.

Pertussis is a complex disease characterised by dynamics that are not well understood, and that can have competing effects in the model [12,14,25–27]. For example, when vaccination coverage decreases, the population who are susceptible to pertussis increases. This can lead to an increase in incidence and transmission. Increases in vaccination decrease transmission and therefore boosting, which may counter-intuitively reduce population protection, especially in older age groups, leading to an increase in incidence and transmission. These competing effects may alter the seroprotection profile of the population and create shifts in the age profile of the burden of disease. These dynamics make it difficult to assess the role of vaccination in reducing pertussis incidence and transmission at the population level. However, as the severity profile is skewed towards infants, vaccination is important for protecting the most vulnerable, even if there is the potential for an increase in total incidence through less severe and/or asymptomatic cases.

A substantial limitation for pertussis models is the lack of contemporary, population-relevant data for parameterisation and a limited understanding of key dynamics [72]. Further, with pertussis lacking a well-defined serological correlate of protection [46], the use of seroprevalence studies to address gaps in available data is not a straightforward option. This presents challenges for model validation and calibration. While our model captures important pertussis dynamics and includes recommended model features, calibrating to a local setting in the absence of reliable, population-wide data on clinical burden presents difficulties for model application and interpretation. The use of modelled burden estimates such as the GBD, or sentinel surveillance data, provide alternative methods for calibration, but both have limitations. One of the approaches proposed for pertussis is to sample the parameter space using LHS and select the parameter sets that capture key dynamics [12]. This alternative to traditional data fitting methods helps mitigate the challenges of using modelled estimates of burden, which fail to capture variation in age and clinical presentation, or reported data, which are usually substantially underreported. Ultimately, model results should be interpretated in light of the complexities of pertussis epidemiology and surveillance.

## 5. Conclusion

We have developed a novel age-structured dynamic, compartmental model of pertussis transmission and vaccination. It explicitly models vaccination, separating each dose of the primary series, booster doses, and maternal vaccination to capture dose-specific effectiveness and duration of protection. We capture heterogeneous immunity profiles and types of infection, with implications for onward transmission. The immune dynamics follow the patterns described in literature, including immune boosting, although the exact characterisation of immunity and the link between infection and disease still have gaps in the literature.

## Data Availability

All relevant data are publicly available or contained within the manuscript and appendices.

## Acknowledgements

We would like to extend our thanks to the Technical Expert Group who served as scientific advisors and provided input on model development, epidemiology and immunology of disease: Dr Ampeire Immaculate, Ugandan National Immunization Program, Ministry of Health, Government of Uganda; Dr Anna Acosta, Meningitis and Vaccine Preventable Diseases Branch at U.S. Centers for Disease Control and Prevention (CDC); Dr Annet Kisakye, Expanded Program on Immunization, World Health Organization, Uganda; Prof Paula Mendes Luz, Instituto Nacional de Infectologia Evandro Chagas (INI); Dr Todi Mengistu, Measurement, Evaluation and Learning (MEL) team at Gavi, the Vaccine Alliance; Dr Helen Quinn, National Centre for Immunisation Research and Surveillance (NCIRS); University of Sydney Children’s Hospital Westmead Clinical School; So Yoon Sim, Value of Vaccines, Modeling & Economics (VoV) team, Immunization Analysis & Insights (IAI) unit, Department of Immunization, Vaccines and Biologicals (IVB), World Health Organization; Dr Rania Tohme, Hepatitis B and Tetanus Team in the Global Immunization Division at the U.S. Centers for Disease Control and Prevention (CDC); and Prof Rudzani Muloiwa, Vaccines for Africa Initiative (VACFA), Faculty of Health Sciences, University of Cape Town, South Africa. We would also like to extend our thanks to Mieke du Plessis, for her valuable support proofreading the manuscript.

## Abbreviations

aP: Acellular pertussis vaccine
CDC: U.S. Centers for Disease Control and Prevention
DTaP: Diphtheria-tetanus-cellular pertussis vaccine
DTP: Diphtheria, tetanus, pertussis
DTPCV: Diphtheria, tetanus, and pertussis-containing vaccine
DTwP: Diphtheria, tetanus and whole-cell pertussis vaccine
EC: Early childhood booster (scenario abbreviation)
EPI: Expanded programme on immunisation
GBD: Global burden of disease
IgG: Immunoglobulin G
LHS: Latin hypercube sampling
LMIC: Low-and middle-income countries
NNDSS: National Notifiable Diseases Surveillance System
PCR: Polymerase chain reaction
PRCC: Partial rank correlation coefficient
PT: Pertussis toxin
SA: Sensitivity analysis
TdaP: Tetanus, diphtheria, acellular pertussis booster dose
TEG: Technical expert group
USA: United States of America
WHO: World Health Organisation
wP: Whole-cell pertussis vaccine

## Appendices – Pertussis Model Details

### Pertussis models

Table A.1 provides a summary of the pertussis models and key features from Campbell et al.’s review, plus four models published since the review was released (indicated with *).

**Table A.1.**
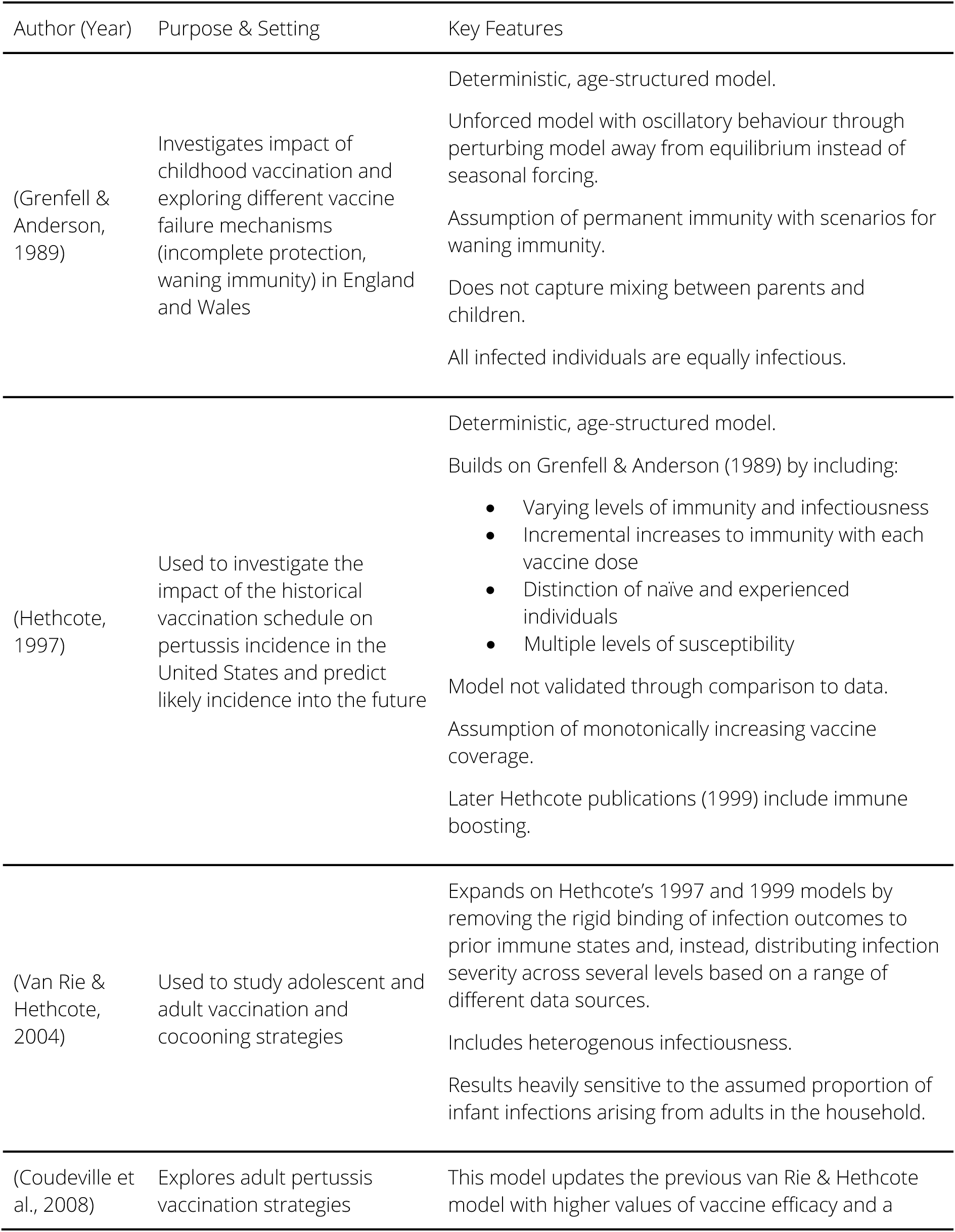
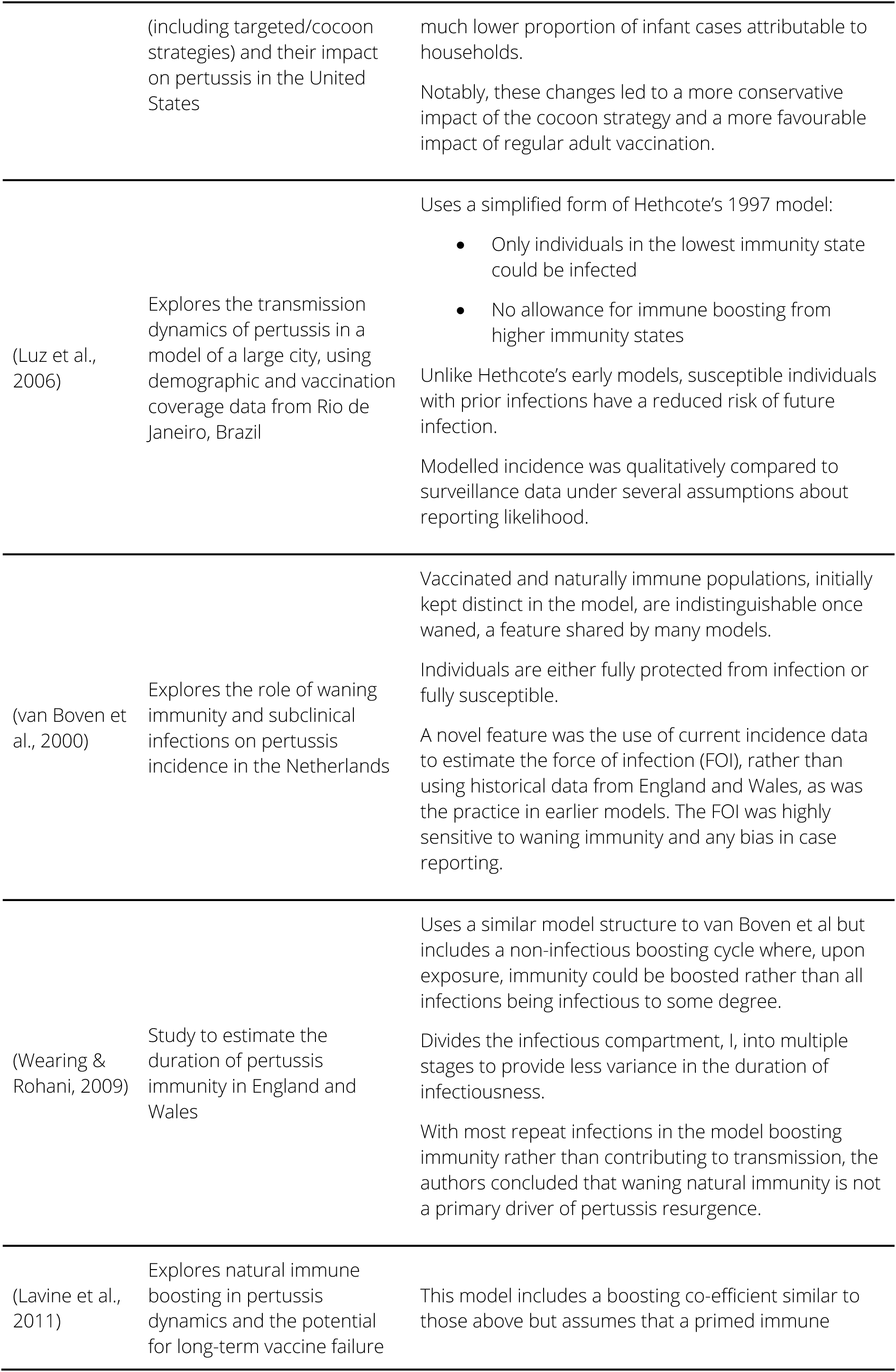
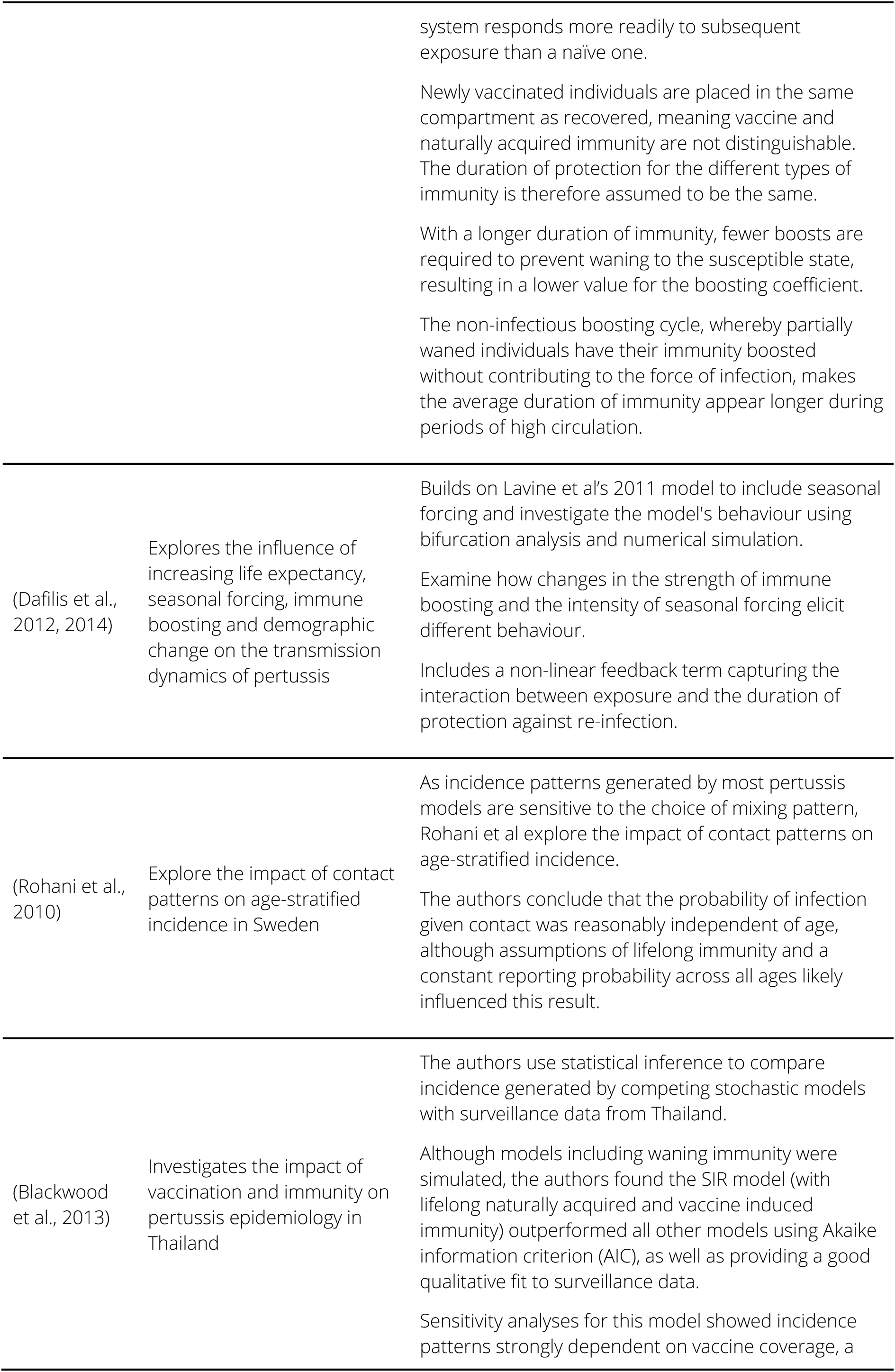
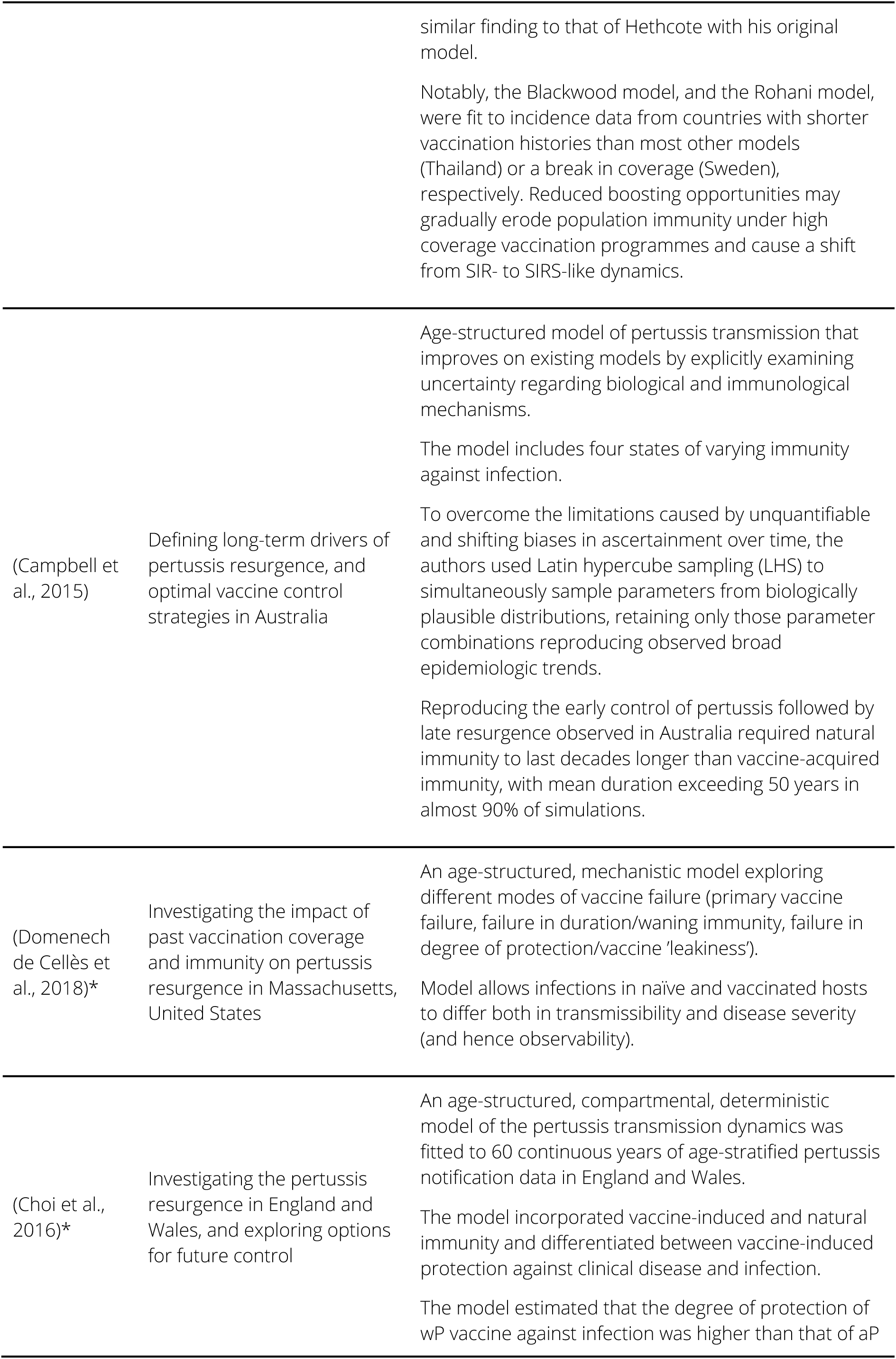
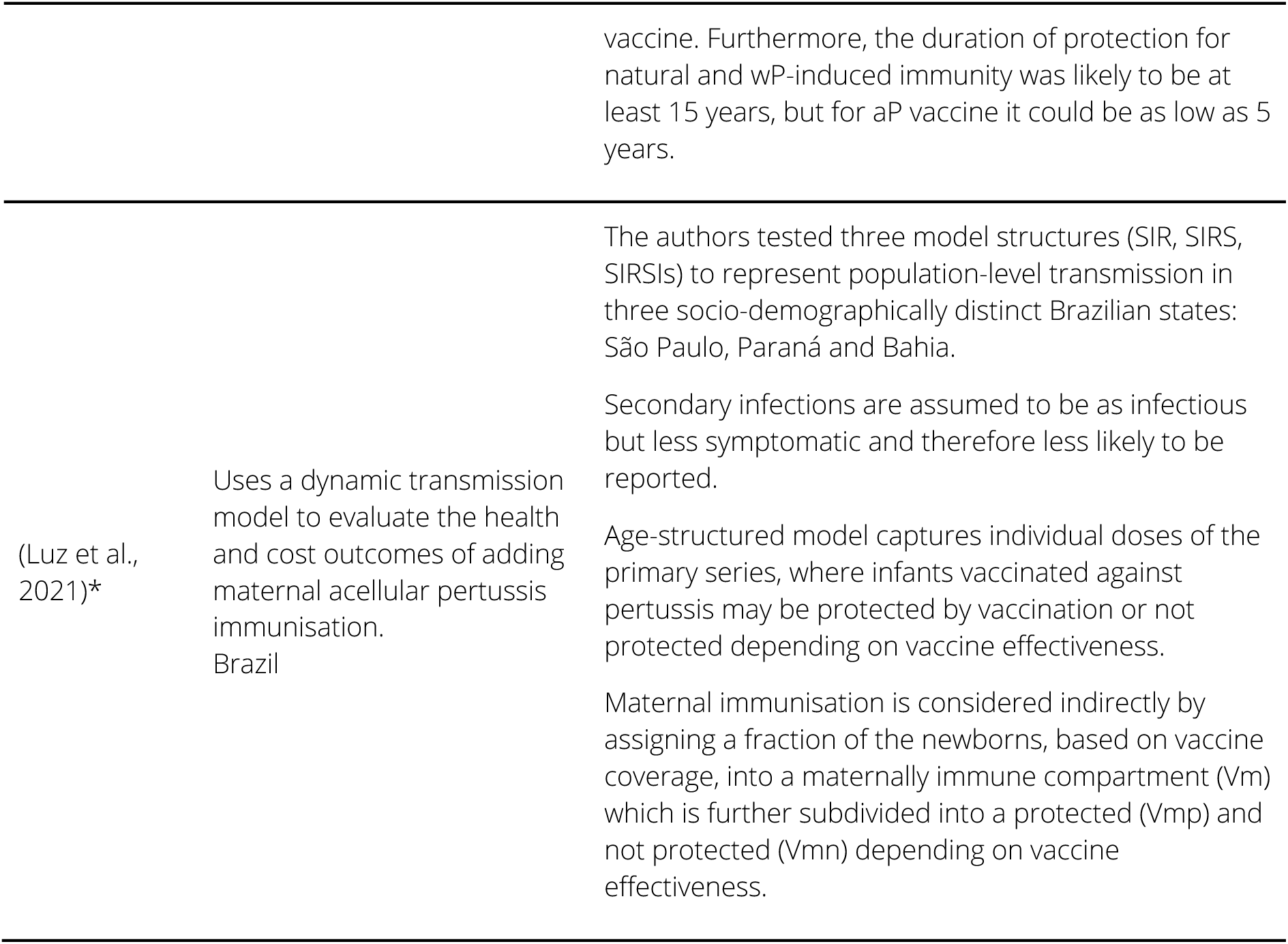
Summary of key pertussis models.

| Author (Year) | Purpose & Setting | Key Features |
| --- | --- | --- |
| (Grenfell & Anderson, 1989) | Investigates impact of childhood vaccination and exploring different vaccine failure mechanisms (incomplete protection, waning immunity) in England and Wales | Deterministic, age-structured model.<br>Unforced model with oscillatory behaviour through perturbing model away from equilibrium instead of seasonal forcing.<br>Assumption of permanent immunity with scenarios for waning immunity.<br>Does not capture mixing between parents and children.<br>All infected individuals are equally infectious. |
| (Hethcote, 1997) | Used to investigate the impact of the historical vaccination schedule on pertussis incidence in the United States and predict likely incidence into the future | Deterministic, age-structured model.<br>Builds on Grenfell & Anderson (1989) by including:<br>• Varying levels of immunity and infectiousness<br>• Incremental increases to immunity with each vaccine dose<br>• Distinction of naïve and experienced individuals<br>• Multiple levels of susceptibility<br>Model not validated through comparison to data.<br>Assumption of monotonically increasing vaccine coverage.<br>Later Hethcote publications (1999) include immune boosting. |
| (Van Rie & Hethcote, 2004) | Used to study adolescent and adult vaccination and cocooning strategies | Expands on Hethcote's 1997 and 1999 models by removing the rigid binding of infection outcomes to prior immune states and, instead, distributing infection severity across several levels based on a range of different data sources.<br>Includes heterogenous infectiousness.<br>Results heavily sensitive to the assumed proportion of infant infections arising from adults in the household. |
| (Coudeville et al., 2008) | Explores adult pertussis vaccination strategies | This model updates the previous van Rie & Hethcote model with higher values of vaccine efficacy and a |
|  | (including targeted/cocoon strategies) and their impact on pertussis in the United States | much lower proportion of infant cases attributable to households.<br>Notably, these changes led to a more conservative impact of the cocoon strategy and a more favourable impact of regular adult vaccination. |
| (Luz et al., 2006) | Explores the transmission dynamics of pertussis in a model of a large city, using demographic and vaccination coverage data from Rio de Janeiro, Brazil | Uses a simplified form of Hethcote's 1997 model:<br>• Only individuals in the lowest immunity state could be infected<br>• No allowance for immune boosting from higher immunity states<br>Unlike Hethcote's early models, susceptible individuals with prior infections have a reduced risk of future infection.<br>Modelled incidence was qualitatively compared to surveillance data under several assumptions about reporting likelihood. |
| (van Boven et al., 2000) | Explores the role of waning immunity and subclinical infections on pertussis incidence in the Netherlands | Vaccinated and naturally immune populations, initially kept distinct in the model, are indistinguishable once waned, a feature shared by many models.<br>Individuals are either fully protected from infection or fully susceptible.<br>A novel feature was the use of current incidence data to estimate the force of infection (FOI), rather than using historical data from England and Wales, as was the practice in earlier models. The FOI was highly sensitive to waning immunity and any bias in case reporting. |
| (Wearing & Rohani, 2009) | Study to estimate the duration of pertussis immunity in England and Wales | Uses a similar model structure to van Boven et al but includes a non-infectious boosting cycle where, upon exposure, immunity could be boosted rather than all infections being infectious to some degree.<br>Divides the infectious compartment, I, into multiple stages to provide less variance in the duration of infectiousness.<br>With most repeat infections in the model boosting immunity rather than contributing to transmission, the authors concluded that waning natural immunity is not a primary driver of pertussis resurgence. |
| (Lavine et al., 2011) | Explores natural immune boosting in pertussis dynamics and the potential for long-term vaccine failure | This model includes a boosting co-efficient similar to those above but assumes that a primed immune |
|  |  | system responds more readily to subsequent exposure than a naïve one.<br>Newly vaccinated individuals are placed in the same compartment as recovered, meaning vaccine and naturally acquired immunity are not distinguishable. The duration of protection for the different types of immunity is therefore assumed to be the same.<br>With a longer duration of immunity, fewer boosts are required to prevent waning to the susceptible state, resulting in a lower value for the boosting coefficient.<br>The non-infectious boosting cycle, whereby partially waned individuals have their immunity boosted without contributing to the force of infection, makes the average duration of immunity appear longer during periods of high circulation. |
| (Dafilis et al., 2012, 2014) | Explores the influence of increasing life expectancy, seasonal forcing, immune boosting and demographic change on the transmission dynamics of pertussis | Builds on Lavine et al's 2011 model to include seasonal forcing and investigate the model's behaviour using bifurcation analysis and numerical simulation.<br>Examine how changes in the strength of immune boosting and the intensity of seasonal forcing elicit different behaviour.<br>Includes a non-linear feedback term capturing the interaction between exposure and the duration of protection against re-infection. |
| (Rohani et al., 2010) | Explore the impact of contact patterns on age-stratified incidence in Sweden | As incidence patterns generated by most pertussis models are sensitive to the choice of mixing pattern, Rohani et al explore the impact of contact patterns on age-stratified incidence.<br>The authors conclude that the probability of infection given contact was reasonably independent of age, although assumptions of lifelong immunity and a constant reporting probability across all ages likely influenced this result. |
| (Blackwood et al., 2013) | Investigates the impact of vaccination and immunity on pertussis epidemiology in Thailand | The authors use statistical inference to compare incidence generated by competing stochastic models with surveillance data from Thailand.<br>Although models including waning immunity were simulated, the authors found the SIR model (with lifelong naturally acquired and vaccine induced immunity) outperformed all other models using Akaike information criterion (AIC), as well as providing a good qualitative fit to surveillance data.<br>Sensitivity analyses for this model showed incidence patterns strongly dependent on vaccine coverage, a |
|  |  | similar finding to that of Hethcote with his original model.<br>Notably, the Blackwood model, and the Rohani model, were fit to incidence data from countries with shorter vaccination histories than most other models (Thailand) or a break in coverage (Sweden), respectively. Reduced boosting opportunities may gradually erode population immunity under high coverage vaccination programmes and cause a shift from SIR- to SIRS-like dynamics. |
| (Campbell et al., 2015) | Defining long-term drivers of pertussis resurgence, and optimal vaccine control strategies in Australia | Age-structured model of pertussis transmission that improves on existing models by explicitly examining uncertainty regarding biological and immunological mechanisms.<br>The model includes four states of varying immunity against infection.<br>To overcome the limitations caused by unquantifiable and shifting biases in ascertainment over time, the authors used Latin hypercube sampling (LHS) to simultaneously sample parameters from biologically plausible distributions, retaining only those parameter combinations reproducing observed broad epidemiologic trends.<br>Reproducing the early control of pertussis followed by late resurgence observed in Australia required natural immunity to last decades longer than vaccine-acquired immunity, with mean duration exceeding 50 years in almost 90% of simulations. |
| (Domenech de Cellès et al., 2018)* | Investigating the impact of past vaccination coverage and immunity on pertussis resurgence in Massachusetts, United States | An age-structured, mechanistic model exploring different modes of vaccine failure (primary vaccine failure, failure in duration/waning immunity, failure in degree of protection/vaccine 'leakiness').<br>Model allows infections in naïve and vaccinated hosts to differ both in transmissibility and disease severity (and hence observability). |
| (Choi et al., 2016)* | Investigating the pertussis resurgence in England and Wales, and exploring options for future control | An age-structured, compartmental, deterministic model of the pertussis transmission dynamics was fitted to 60 continuous years of age-stratified pertussis notification data in England and Wales.<br>The model incorporated vaccine-induced and natural immunity and differentiated between vaccine-induced protection against clinical disease and infection.<br>The model estimated that the degree of protection of wP vaccine against infection was higher than that of aP |
|  |  | vaccine. Furthermore, the duration of protection for natural and wP-induced immunity was likely to be at least 15 years, but for aP vaccine it could be as low as 5 years. |
| (Luz et al., 2021)* | Uses a dynamic transmission model to evaluate the health and cost outcomes of adding maternal acellular pertussis immunisation. Brazil | The authors tested three model structures (SIR, SIRS, SIRSIs) to represent population-level transmission in three socio-demographically distinct Brazilian states: São Paulo, Paraná and Bahia.<br>Secondary infections are assumed to be as infectious but less symptomatic and therefore less likely to be reported.<br>Age-structured model captures individual doses of the primary series, where infants vaccinated against pertussis may be protected by vaccination or not protected depending on vaccine effectiveness.<br>Maternal immunisation is considered indirectly by assigning a fraction of the newborns, based on vaccine coverage, into a maternally immune compartment ($V_m$) which is further subdivided into a protected ($V_{mp}$) and not protected ($V_{mn}$) depending on vaccine effectiveness. |

### Age groups

**Table A.2.**
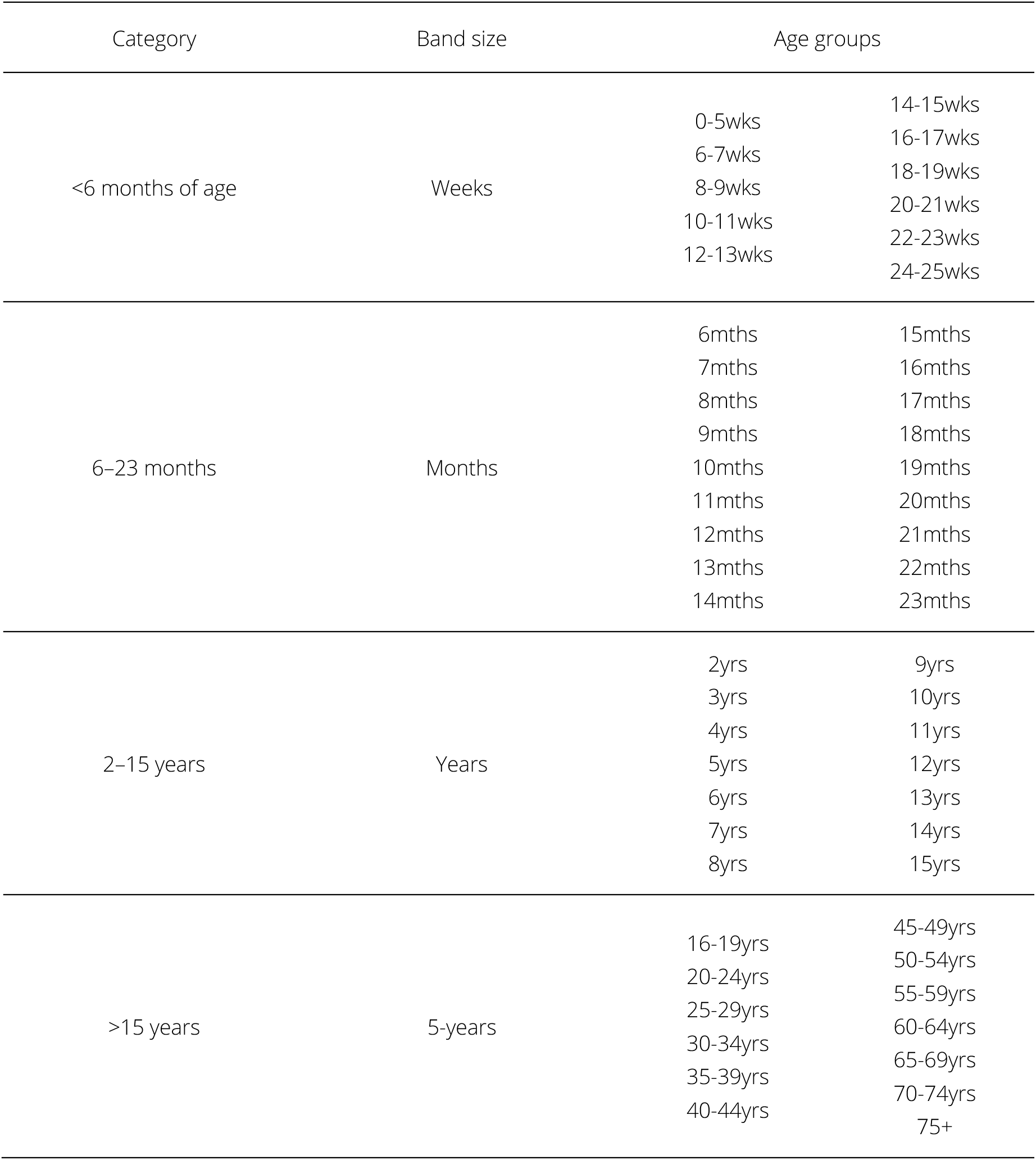
Model age groups.

### Model structure

**Table A.3.**
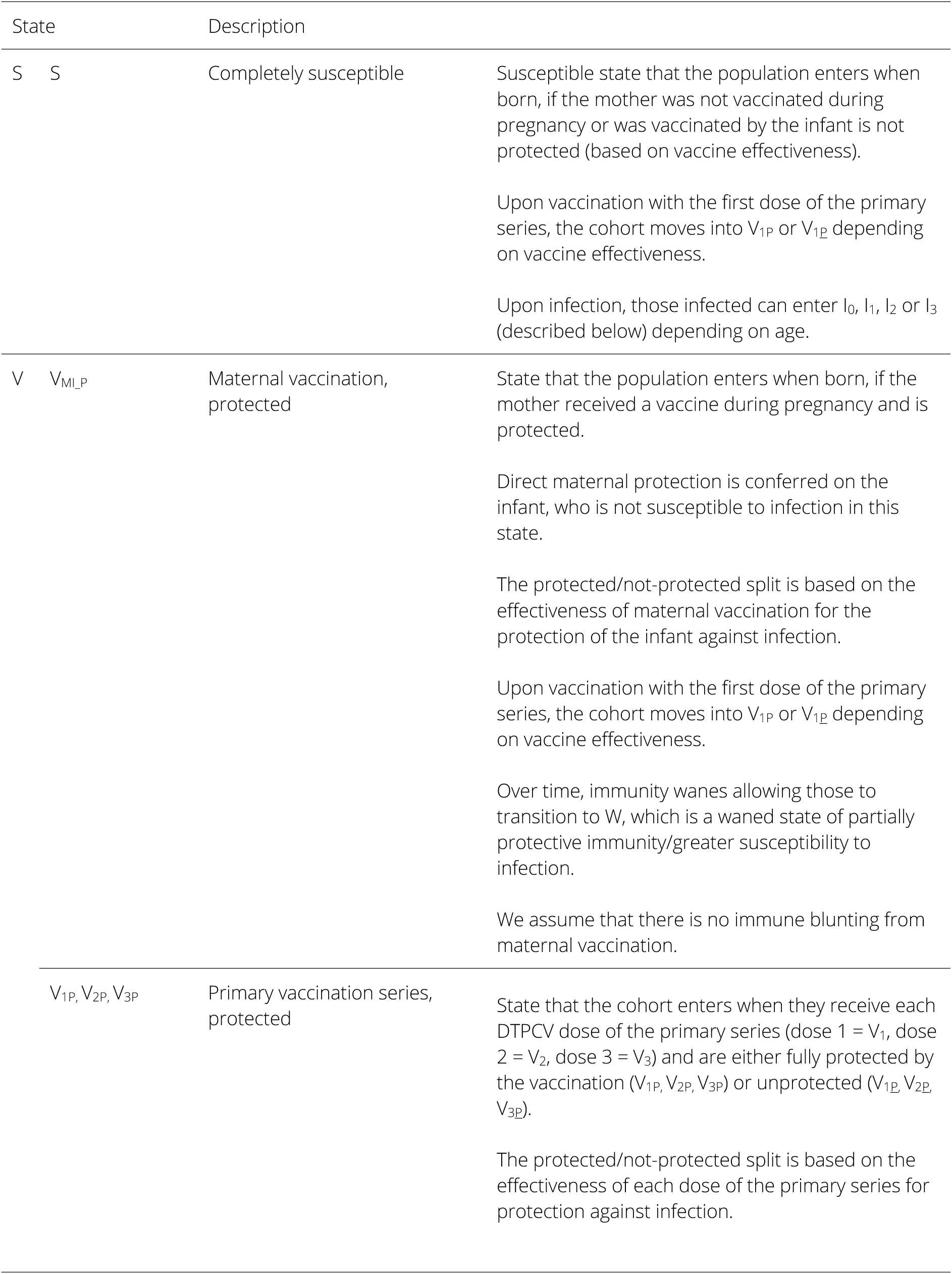
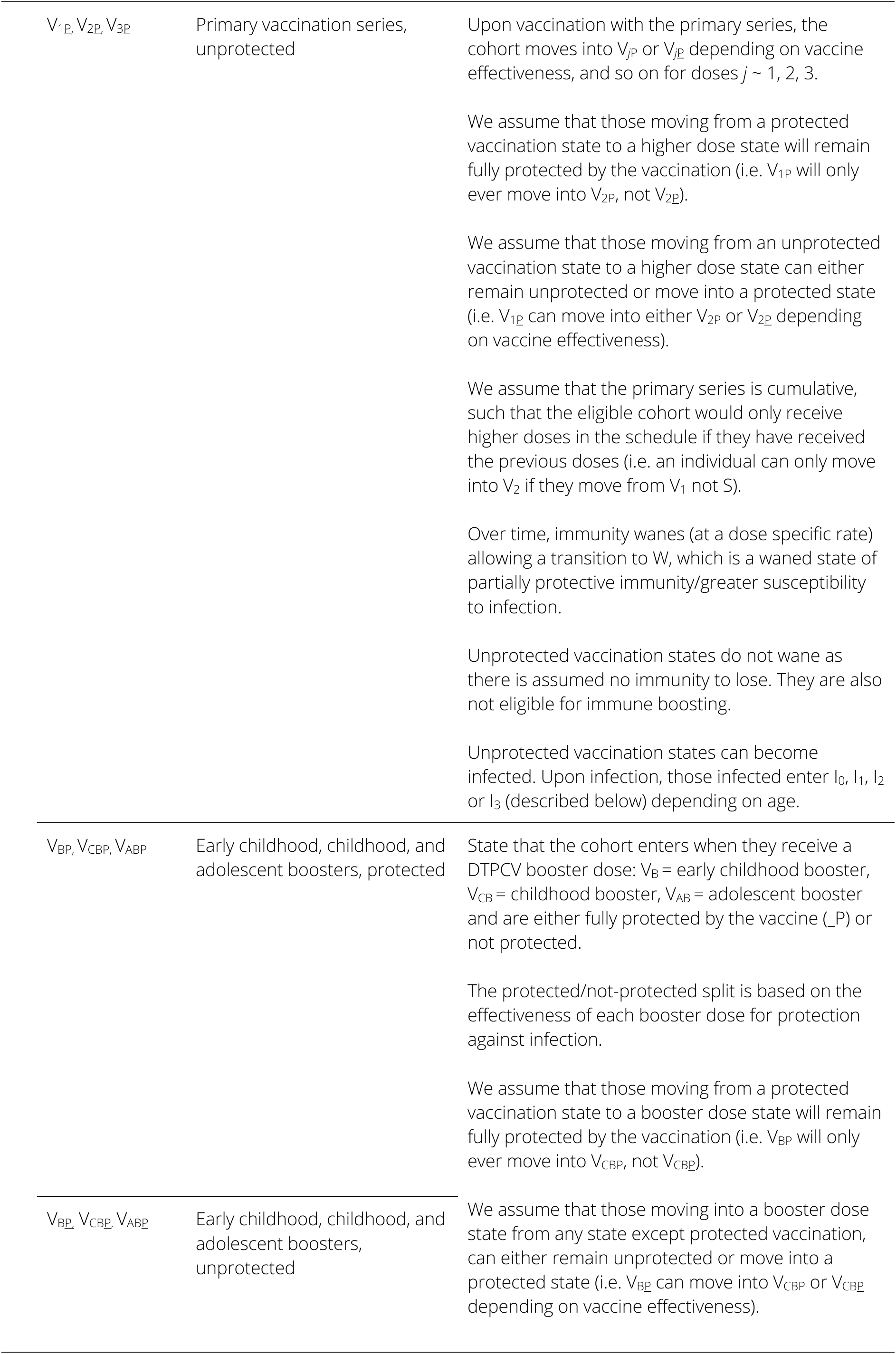
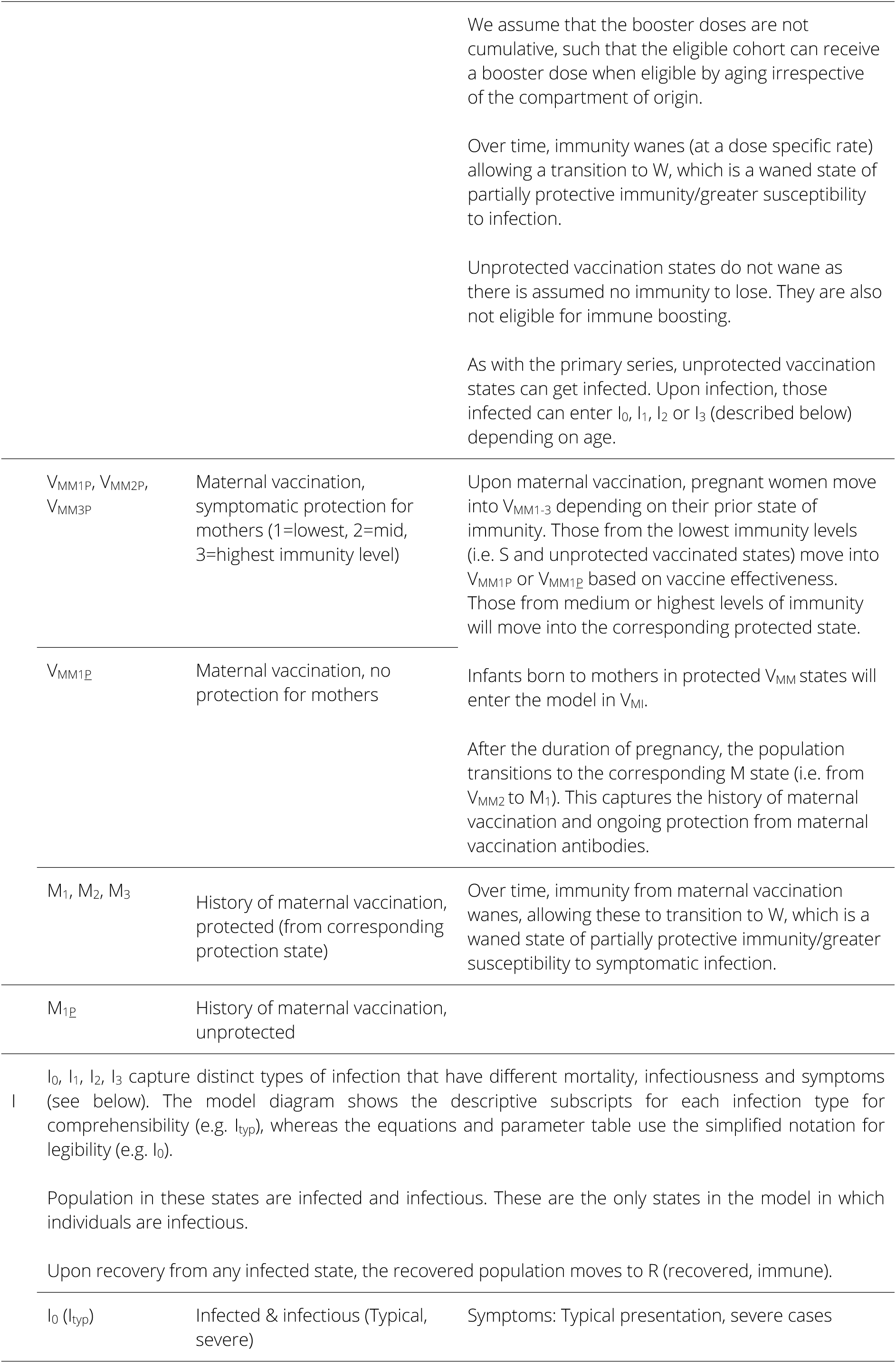
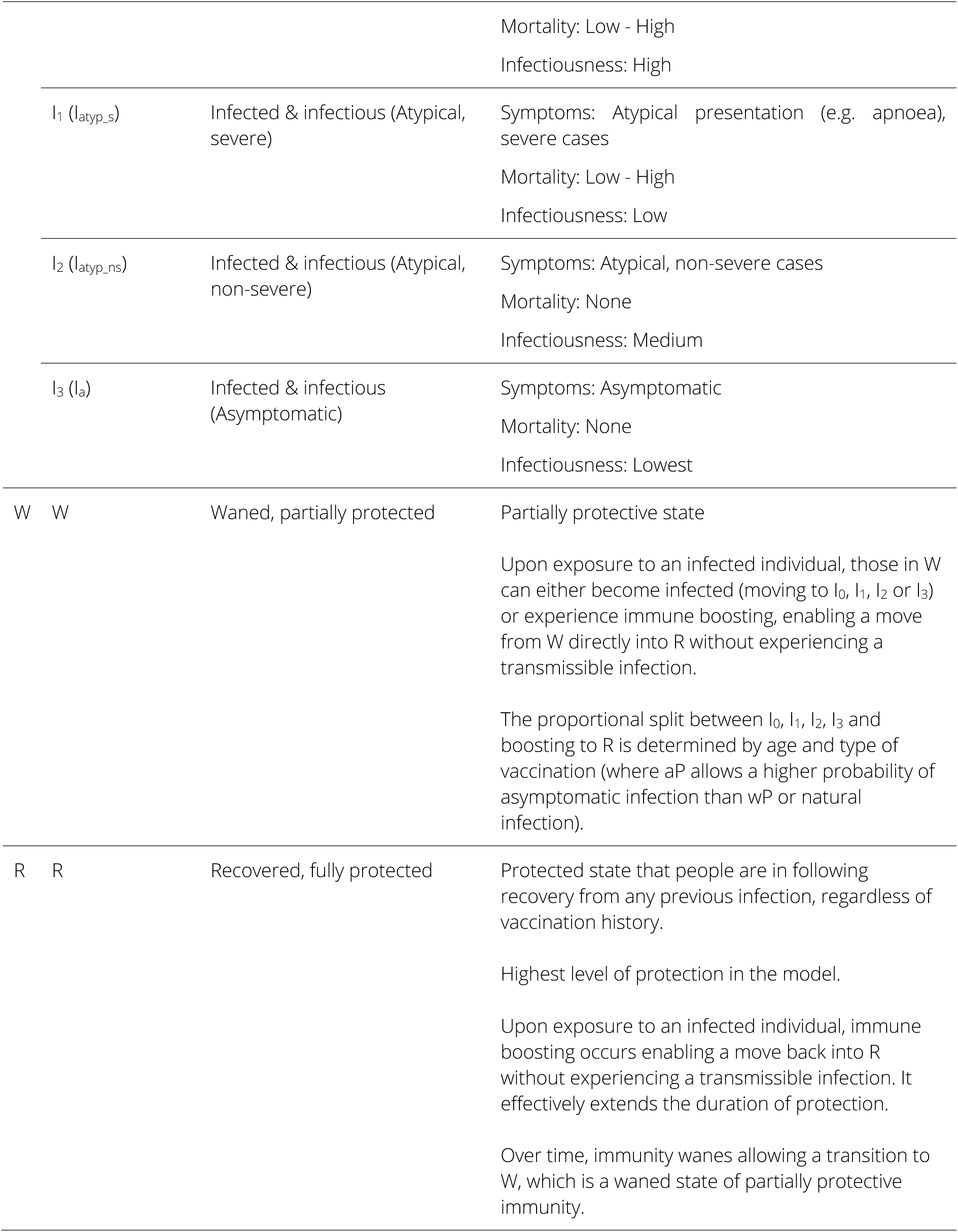
Pertussis model compartments (states), descriptions and key assumptions.

### Model equations

The rates of change of the population in each epidemiological state are described by the following set of ordinary differential equations, for each age group *i* = 56. For simplicity, the notation for the infectious states uses the condensed format (I_0_, I_1_, I_2_, I_3_) detailed in Table A.3.

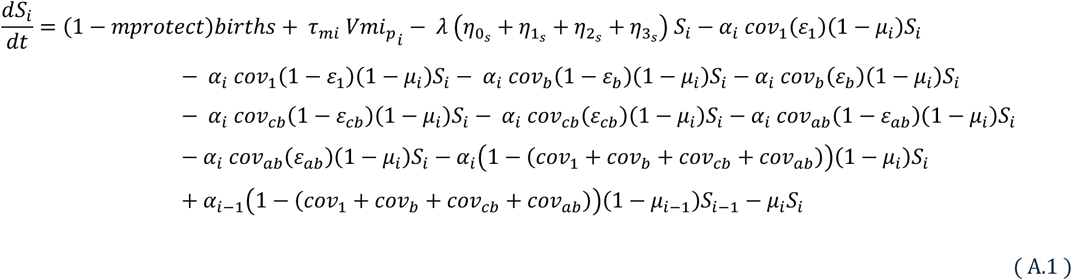

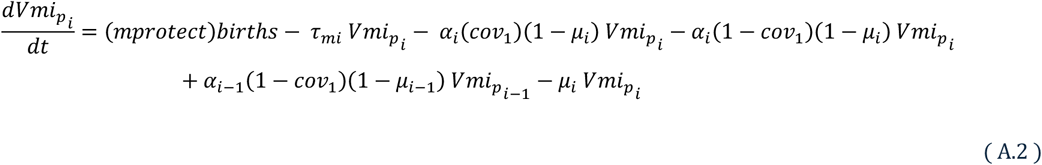

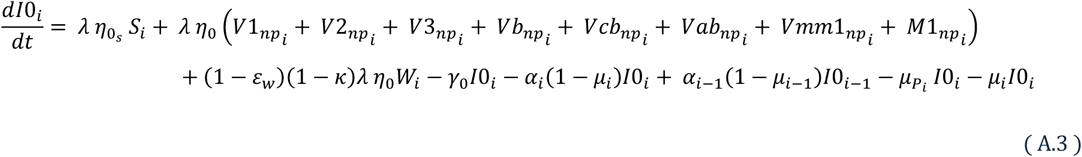

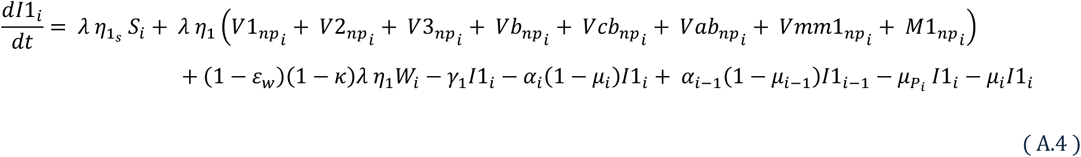

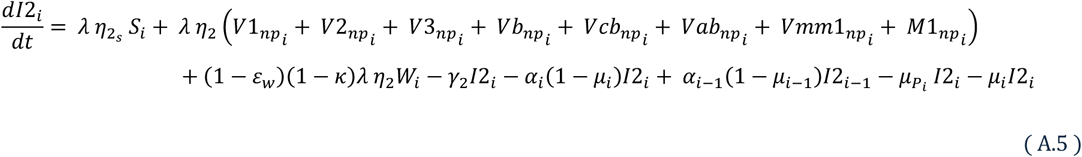

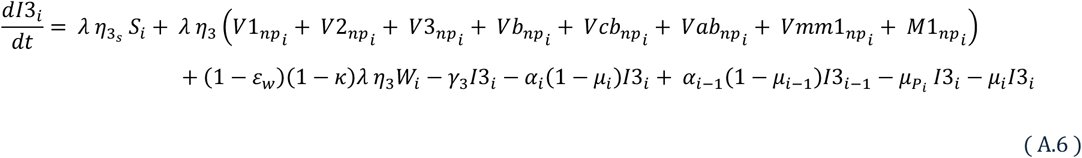

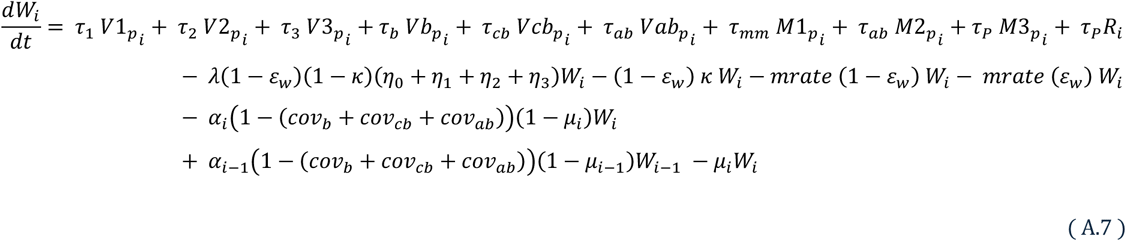

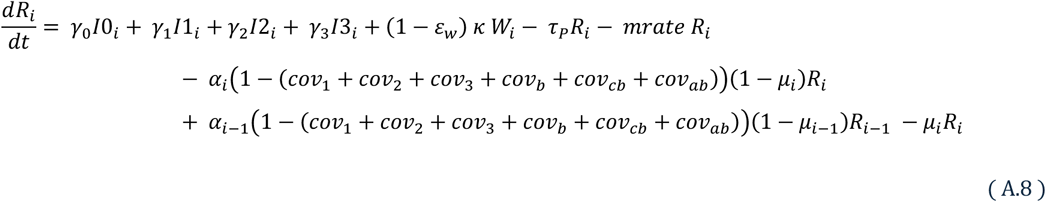

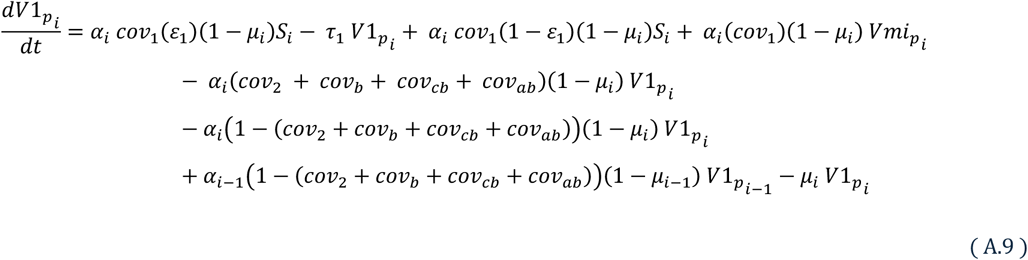

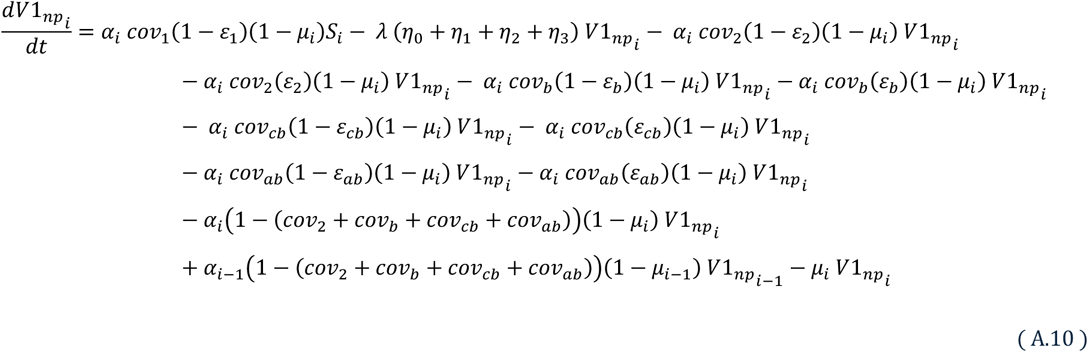

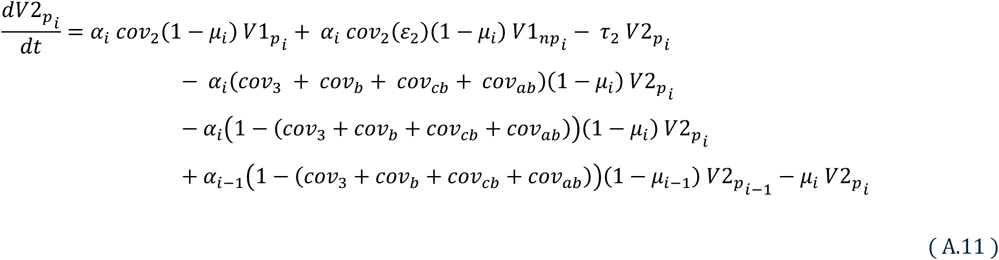

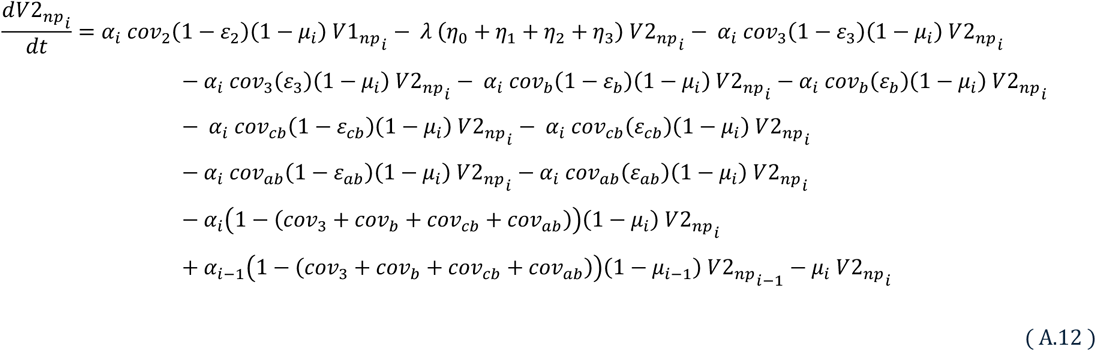

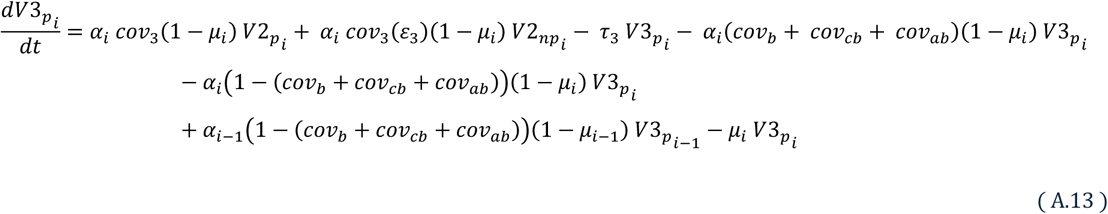

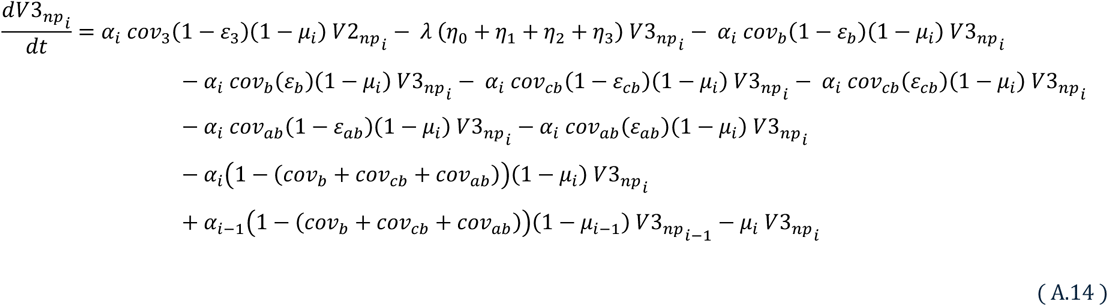

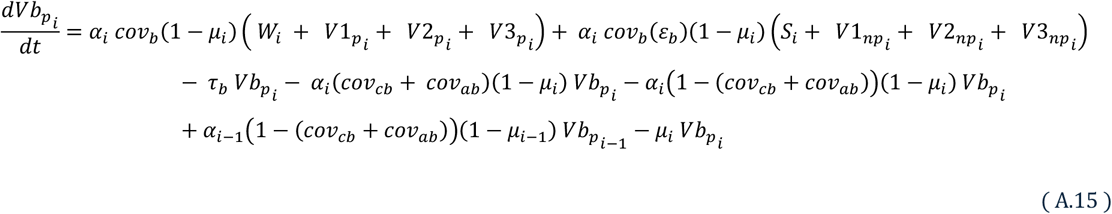

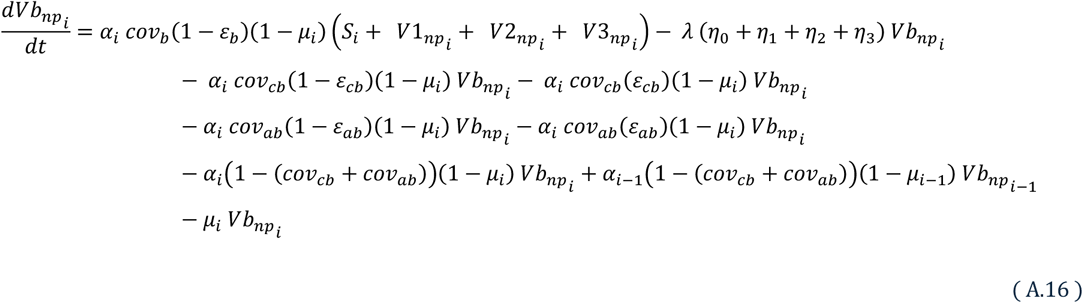

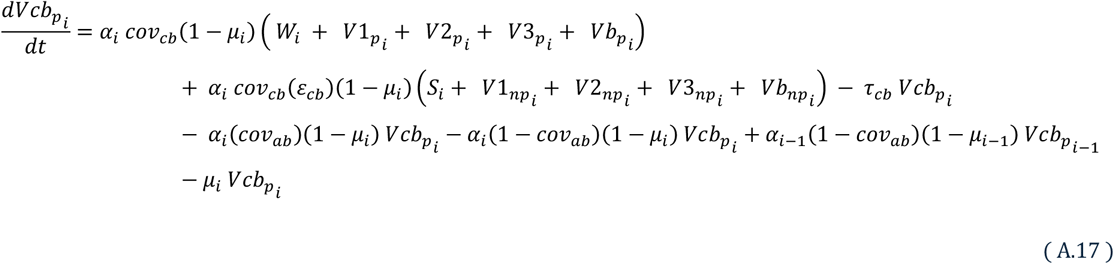

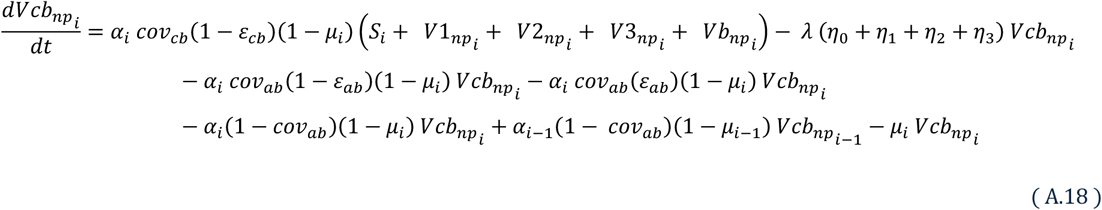

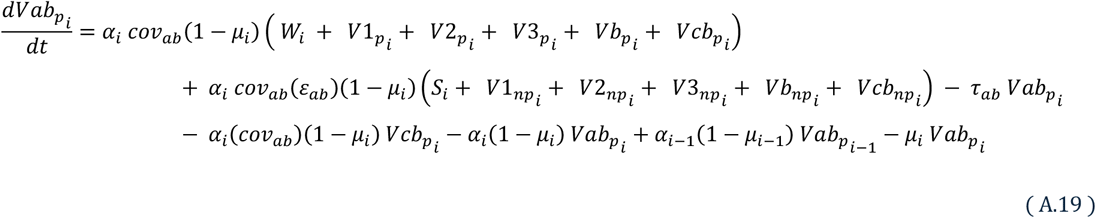

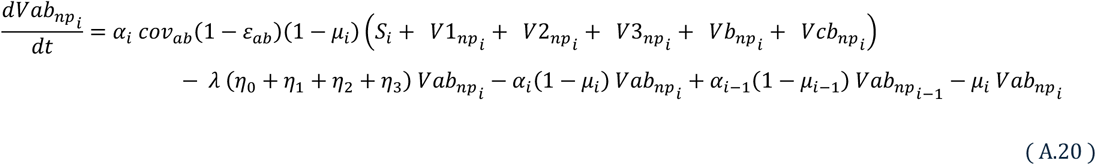

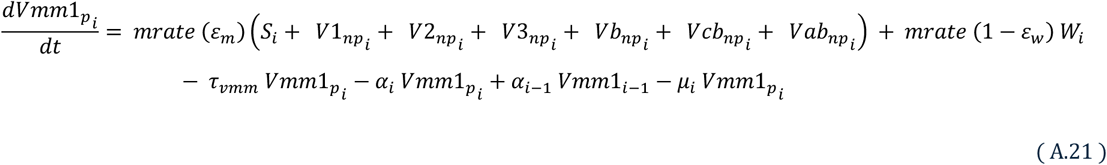

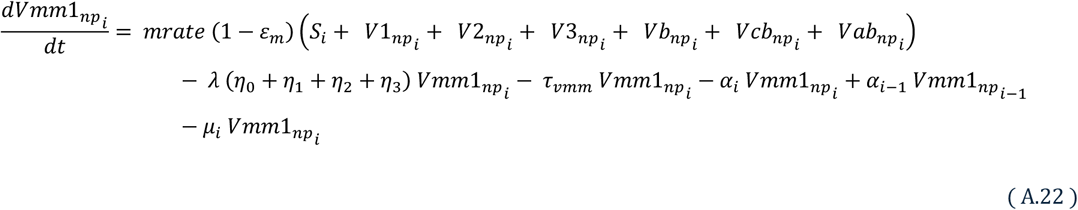

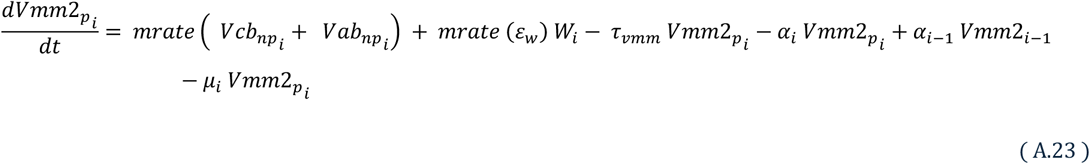

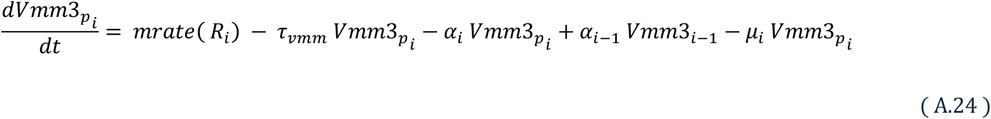

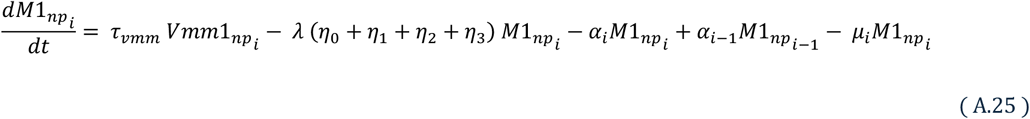

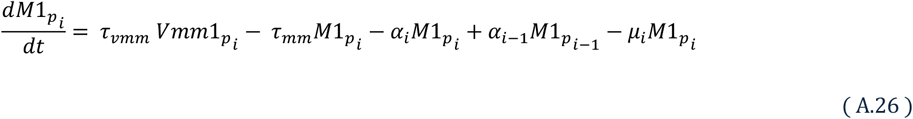

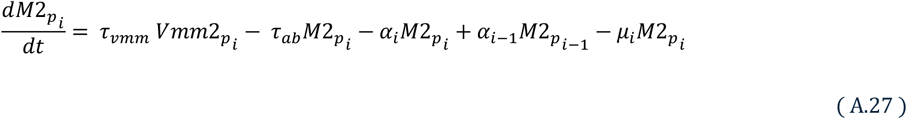

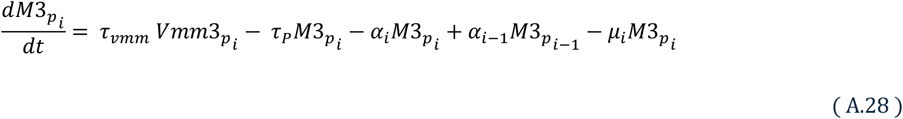

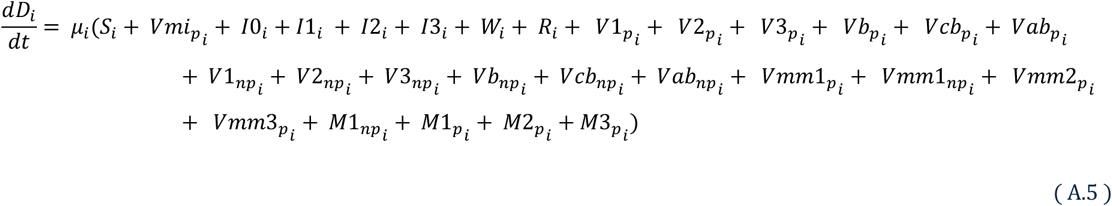

Where λ, the force of infection, is:

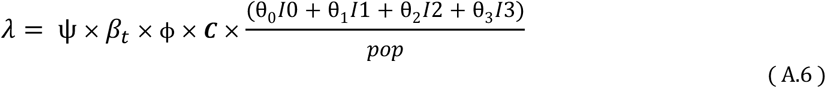

and *pop* is the sum of all compartments at time t, except D (death)

Periodicity is introduced in the force of infection equation, λ, as follows:

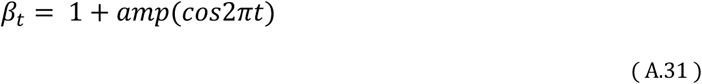

Stochasticity, denoted by 𝜓, is sampled from a uniform distribution and incorporated as a multiplier term in the force of infection equation, λ. Stochasticity captures the probabilistic nature of transmission and demographics. It can be removed from the model for a deterministic run.

### Parameters

**Table A.4.**
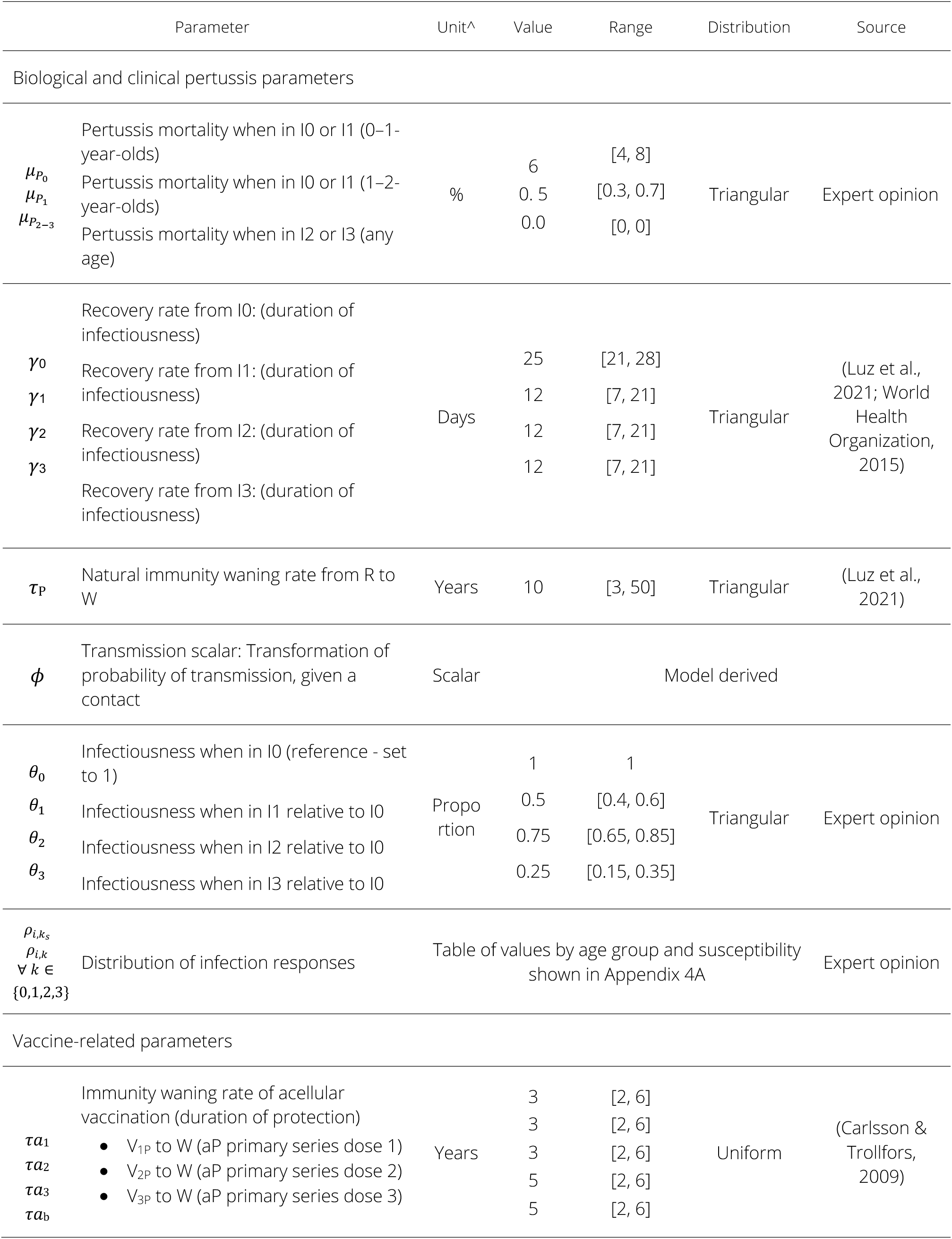
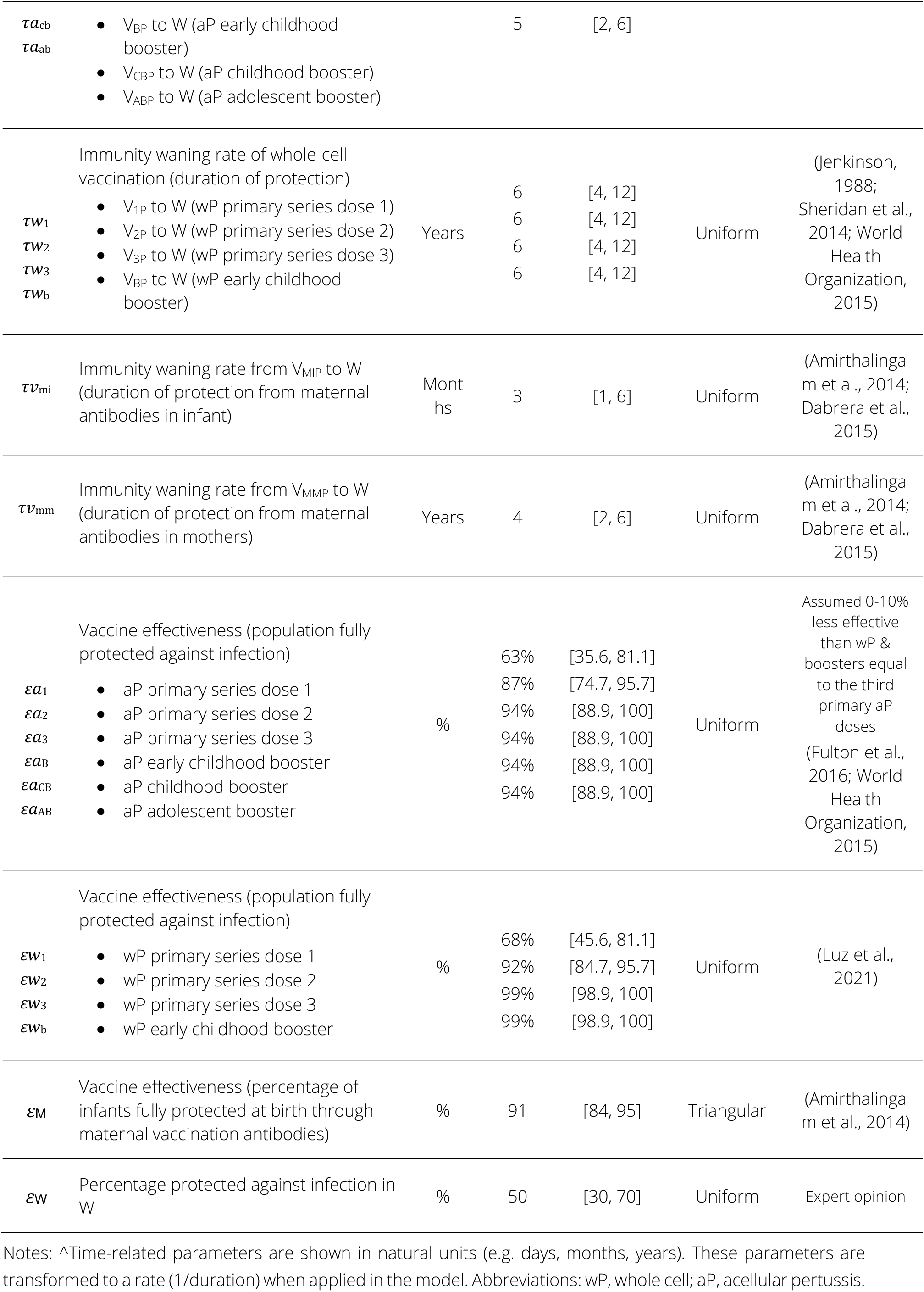
Model parameters values for pertussis.

**Table 4A.1.**
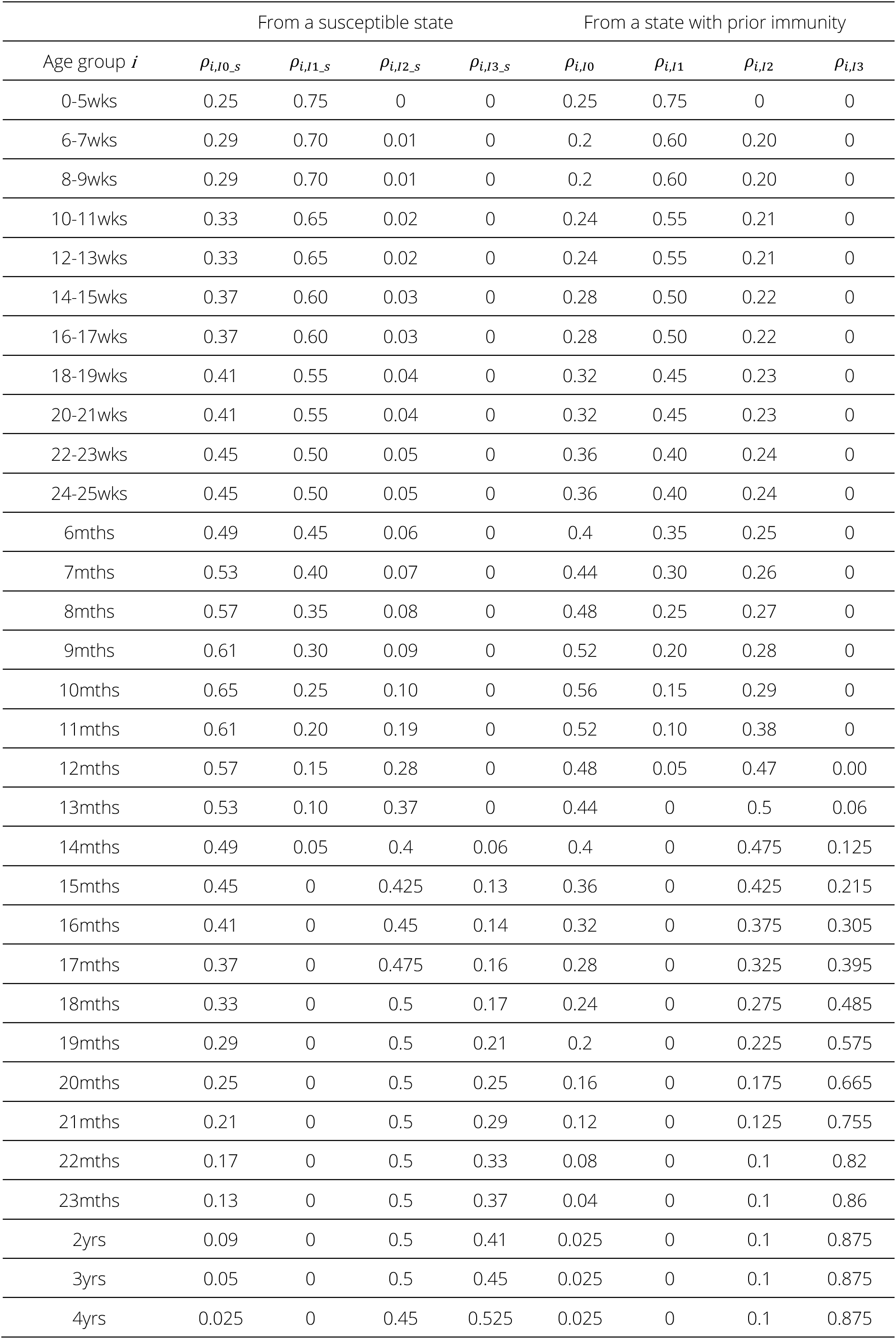
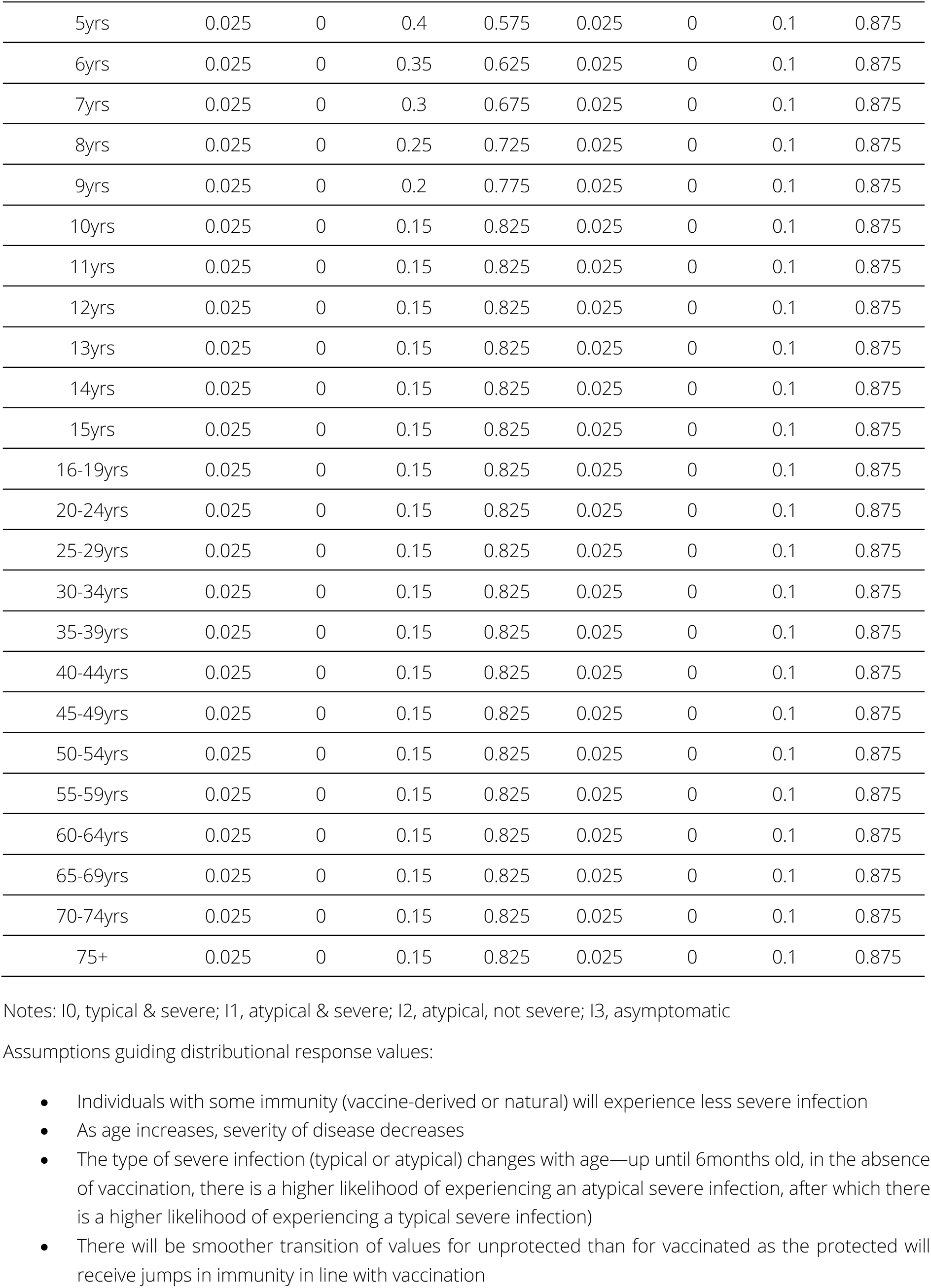
Distribution of infection responses for pertussis.

## Sensitivity analysis

### Sensitivity analysis approach

We conducted a sensitivity analysis (SA) on the pertussis model using Latin hypercube sampling (LHS)-partial rank correlation coefficient (PRCC) (Blower, S.M., 1994; Marino et al., 2008; Wu et al., 2013). Drawing from the appropriate parameter distributions (Table A.4), we conducted LHS of the parameter space to generate 1000 parameter sets. LHS for the immunity waning parameters was drawn from a uniform distribution between 1 and 3 for each set, where 2 represents the default value, 1 the minimum duration of protection, and 3 the maximum duration of protection. As we only explore vaccination impact for the primary series, early childhood booster, and maternal vaccination, and not the childhood and adolescent boosters, we did not separate the doses into distinct groups. Instead, we varied them simultaneously so that the relative duration of protection across doses was maintained.

After running the model 1000 times on each of the LHS parameter sets and obtaining the corresponding model outputs, we calculated the PRCC using the epiR package (version 2.0.80) in R. Input variables for the PRCC are: pertussis transmission scalar; likelihood of death; relative infectiousness of infection types I_1_, I_2_, and I_3_ compared to I_0_; recovery rate of infection types I_0_, I_1_, I_2_, and I_3_; boosting proportion; immunity waning rate (infection); immunity waning rate (vaccine); waned protection; and vaccine effectiveness. Three model outputs (average annual pertussis incidence, proportion of the population protected against pertussis, and average annual number of pertussis deaths) for three age groups (all ages, under 5-year-olds, and under 15-year-olds) are considered for the PRCC.

### Sensitivity analysis findings

Figure A.1 shows the PRCC from 1000 model runs of a scenario with the primary series only (no booster doses). The bars are coloured to make the input variables easily distinguishable across the nine panels. The black lines show the 95% confidence interval (CI) for the PRCC. Where the CI crosses zero, the PRCC for the corresponding input parameter is not statistically significant at the 5% level.

Modelled incidence and deaths are most sensitive to changes in immunity waning rates for naturally acquired and vaccine-derived immunity, the boosting proportion, and the recovery rate of severe atypical infections (I_1_). This is consistent across the three age categories considered. Modelled estimates of population protected are most sensitive to changes in the transmission scalar, immune boosting parameters (boosting proportion and waned protection), and the relative infectiousness of asymptomatic infections, across the three age categories. The sensitivity of population protection to the immune waning rate for vaccine-derived and naturally acquired protection varies by age category. The immunity waning rate of vaccine-derived protection is the third most sensitive parameter in under 5-year-olds, the seventh most sensitive parameter in all ages, and is not significant in the under 15-year-olds.

Transmission scalar: The transmission scalar (the parameter used for calibration), is significant for all three model outputs across all age categories. However, the model is most sensitive to changes in the transmission scalar for population protection (three to seven times larger PRCC than for incidence and deaths). This aligns with expectations as naturally acquired immunity drives a substantial proportion of population protection in the model.

Relative infectiousness: The relative infectiousness of asymptomatic infections (I_3_) is not significant for incidence or deaths but is a significant sensitive parameter for population protection where it is positively correlated for each of the three age categories. This behaviour is to be expected as asymptomatic infections are a large proportion of the total burden. Increases in the relative infectious of asymptomatic cases will raise the total burden of pertussis and drive an increase in naturally acquired immunity.

Immune boosting: The boosting proportion (i.e. the proportion of the population who are boosted from W to R instead of experiencing a transmissible infection, W to I) is a sensitive parameter for all three model outputs. While significant for population protected in each of the three age categories, there is a stronger correlation (i.e. higher sensitivity) in the all-ages category than in under five-year-olds and under 15-year-olds. The correlation is negative for all model outputs in the SA: the more people who experience immune boosting instead of a transmissible infection, the lower the incidence, population protected, and deaths. More boosting decreases the population protected because there is less transmission to drive onward infection, which would result in a direct and indirect effect on population immunity.

Immune waning: Both vaccine-derived and naturally acquired immunity waning rates are highly sensitive parameters. Due to the structure of the LHS, the directionality of the waning rates in the PRCC for infection and vaccination have opposite signs—i.e. for the immunity waning rate (infection), a larger value corresponds to a shorter duration of protection whereas for the immunity waning rate (vaccine), a larger value corresponds to a longer duration of protection. In the model, the parameters are transformed to be consistent with the interpretation of waning, such that a longer duration of protection leads to a smaller immunity waning rate. This makes the PRCC for the two immunity waning rates appear to have opposite impacts on the model outputs of interest. In practice, for both vaccine-derived and naturally acquired immunity, the longer the duration of protection, the smaller the incidence and deaths (consistent across each of the three age categories).

**Figure A.1.**
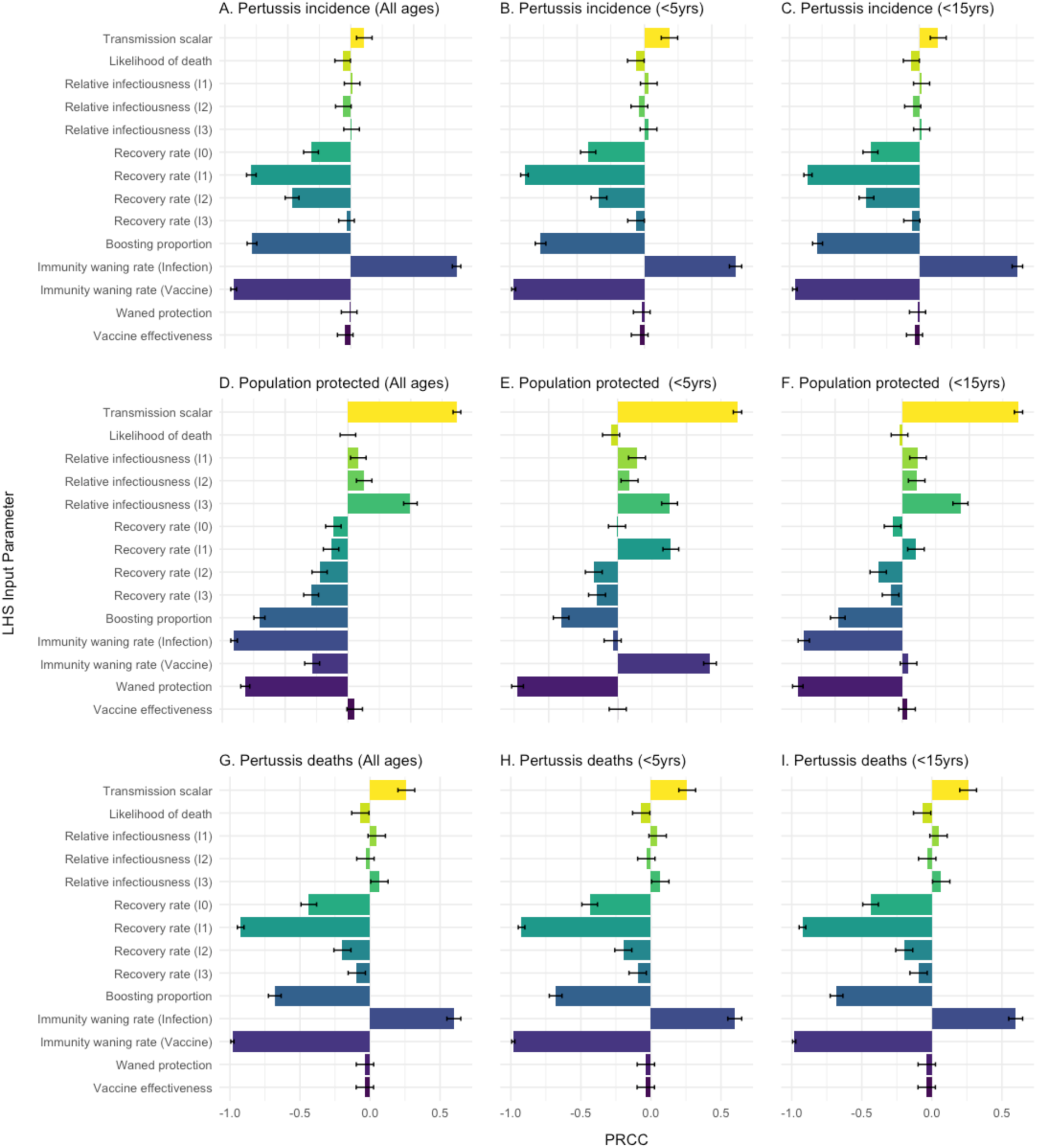
Partial rank correlation coefficient (PRCC) of pertussis parameters in relation to pertussis incidence, population protected, and deaths, for the DTP primary series Figure description: PRCC of pertussis parameters in relation to pertussis incidence (Panels A – C), proportion of the population protected (%) (Panels D – F), and pertussis deaths (Panels G – H). The sensitivity analysis is subset by age: All ages (Panels A, D, G), children under 5-years of age (Panels B, E, H), and children under 15 years of age (Panels C, F, I). PRCC values above zero are positively correlated with the output variable and those below zero are negatively correlated with the output variable. The 95% confidence interval is shown in the black whiskers on each bar. The values are drawn from 1000 model runs. LHS, Latin hypercube sampling, PRCC, Partial rank correlation coefficient.

Figure A.2 shows the relationship between pertussis incidence and the duration of protection from vaccination and infection. There is a clear pattern of increasing pertussis incidence with shorter durations of both vaccine-derived and naturally acquired immunity.

**Figure A.2.**
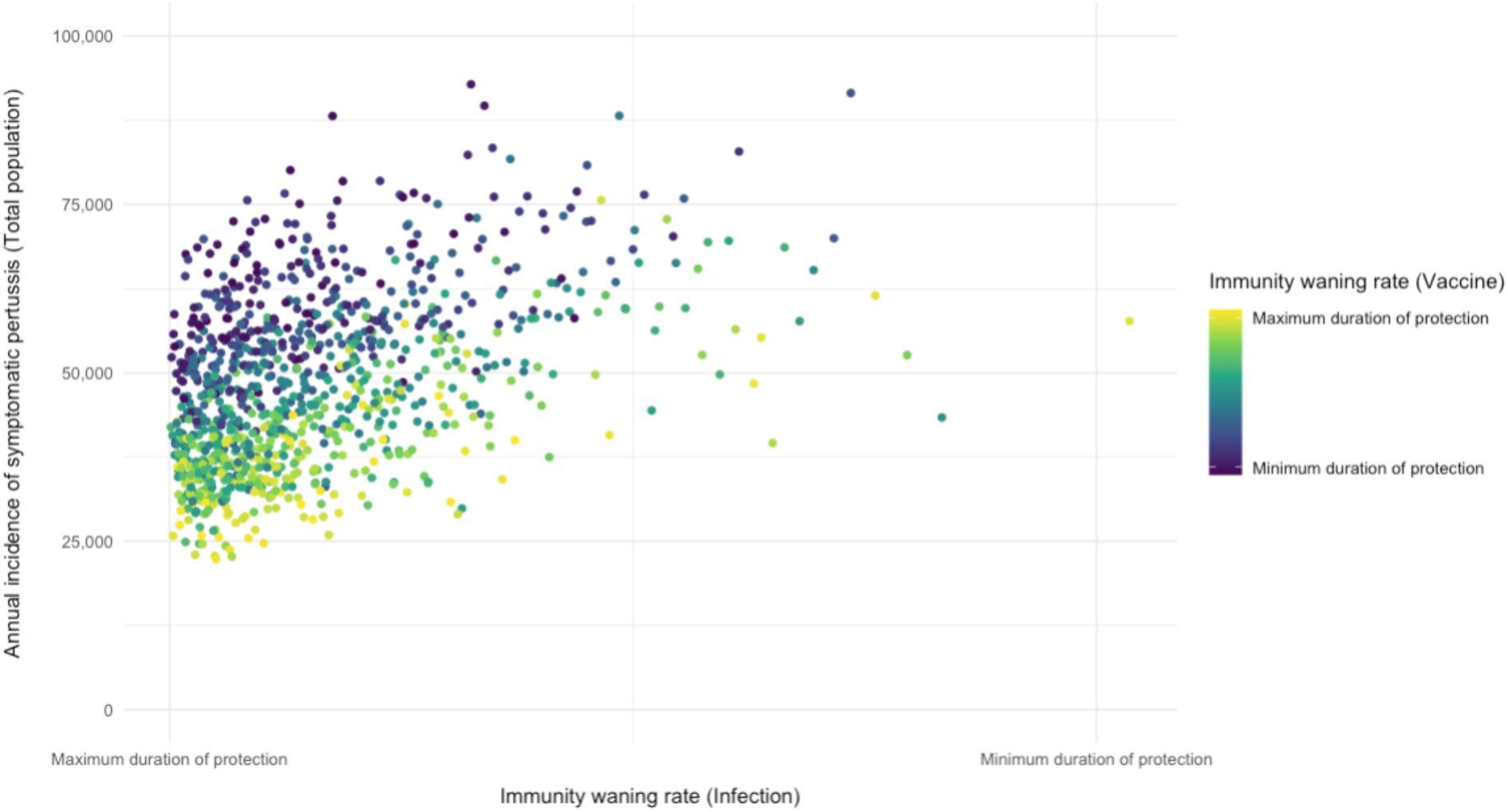
Projected annual incidence of symptomatic pertussis cases in all ages for each of the 1000 LHS parameter sets, shown in relation to the waning rates of naturally acquired and vaccine-derived immunity Figure description: Each point represents a model run with one of the 1000 LHS sets. The points are coloured according to the range of values for the waning rate of vaccine derived immunity (from dark purple representing the minimum modelled value to yellow representing the maximum modelled value for duration of protection against pertussis). Immune waning is 1/duration of protection. See Table S4 for ranges and sources. LHS, Latin hypercube sampling.

### Technical Expert Group

The role of the technical expert group (TEG) was to advise on the model development, epidemiology and immunology of disease, and broader project guidance. The parameter values were established with input and review from the TEG.

● Dr Ampeire Immaculate, Ugandan National Immunization Program, Ministry of Health, Government of Uganda.
● Dr Anna Acosta, Meningitis and Vaccine Preventable Diseases Branch at U.S. Centers for Disease Control and Prevention (CDC)
● Dr Annet Kisakye, Expanded Program on Immunization, World Health Organization, Uganda
● Prof Paula Mendes Luz, Instituto Nacional de Infectologia Evandro Chagas (INI)
● Dr Todi Mengistu, Measurement, Evaluation and Learning (MEL) team at Gavi, the Vaccine Alliance
● Dr Helen Quinn, National Centre for Immunisation Research and Surveillance (NCIRS); University of Sydney Children’s Hospital Westmead Clinical School.
● So Yoon Sim, Value of Vaccines, Modeling & Economics (VoV) team, Immunization Analysis & Insights (IAI) unit, Department of Immunization, Vaccines and Biologicals (IVB), World Health Organization
● Dr Rania Tohme, Hepatitis B and Tetanus Team in the Global Immunization Division at the U.S. Centers for Disease Control and Prevention (CDC)

## Notes

### Competing Interest Statement

The authors have declared no competing interest.

### Funding Statement

This work was supported by the U.S. Centers for Disease Control and Prevention through a cooperative agreement to the African Field Epidemiology Network (AFENET). Rachel Hounsell received funding through the Nuffield Department of Medicine Prize Fellowship at the University of Oxford with support from the Engineering and Physical Sciences Research Council (EPSRC) at UKRI and Green Templeton College.

